# Genome-Wide Association Studies of Coffee Intake in UK/US Participants of European Ancestry Uncover Gene-Cohort Influences

**DOI:** 10.1101/2023.09.09.23295284

**Authors:** Hayley H A Thorpe, Pierre Fontanillas, Benjamin K Pham, John J Meredith, Mariela V Jennings, Natasia S Courchesne-Krak, Laura Vilar-Ribó, Sevim B Bianchi, Julian Mutz, 23andMe Research Team, Sarah L Elson, Jibran Y Khokhar, Abdel Abdellaoui, Lea K Davis, Abraham A Palmer, Sandra Sanchez-Roige

## Abstract

Coffee is one of the most widely consumed beverages. We performed a genome-wide association study (**GWAS**) of coffee intake in US-based 23andMe participants (*N*=130,153) and identified 7 significant loci, with many replicating in three multi-ancestral cohorts. We examined genetic correlations and performed a phenome-wide association study across thousands of biomarkers and health and lifestyle traits, then compared our results to the largest available GWAS of coffee intake from UK Biobank (UKB; *N*=334,659). The results of these two GWAS were highly discrepant. We observed positive genetic correlations between coffee intake and psychiatric illnesses, pain, and gastrointestinal traits in 23andMe that were absent or negative in UKB. Genetic correlations with cognition were negative in 23andMe but positive in UKB. The only consistent observations were positive genetic correlations with substance use and obesity. Our study shows that GWAS in different cohorts could capture cultural differences in the relationship between behavior and genetics.

## Introduction

Coffee is a leading global food commodity that has psychoactive properties that are largely due to the presence of caffeine^1^. While rates of use and daily intake varies widely by geographic region, it is estimated that approximately 60-85% of adults in Europe and the United States consume between 0.6 to 5.5 cups of coffee daily^2–4^. Intake of coffee and its bioactive constituents is associated with benefits on cognitive function^5^ and lower risk of liver disease^6,7^ (but see^8^), Parkinson’s and other neurodegenerative diseases^6,7,9^, cardiovascular disease^6,7^, type II diabetes^6,7^, and certain cancers^6,7,10^. However, coffee intake is also associated with higher risks for some adverse outcomes, including increased risk of other substance use and misuse^11–14^, some cancers (e.g., lung cancer^7,10,15^), poor lipid profile^6,7^, pregnancy loss^6,7^, gastrointestinal maladies^16^, and worse cardiovascular outcomes following excessive intake^17^. Given the widespread and regular intake of coffee across the globe, addressing the full spectrum of correlations with health and disease is an important but challenging task.

Genetic studies offer a compelling avenue to investigate the relationships between coffee intake and other complex traits. Twin studies that calculate genetic contributions to daily coffee intake estimate it to be 36-56% heritable, suggesting that coffee intake should be amenable to genetic analysis. Whereas phenotypic correlations, which depend on measuring two or more traits in the same cohort, can arise from genetic and environmental factors, genetic correlations assess genetically driven relationships using the results from genome-wide association studies (**GWAS**) and can therefore examine correlations between two or more traits, even if they were measured in entirely non-overlapping cohorts. In the past decade, over a dozen GWAS (*N=*1,207-407,072) have examined coffee intake^18–34^. Several of these GWAS have found associations with single nucleotide polymorphisms (**SNPs**) within or near genes that metabolize caffeine (**Supplementary Table 1**), such as *CYP1A1* and *CYP1A2*^18–20,23–26,30,32,33^. Some of these loci are also associated with other complex traits, including liver disease^35–37^, cancers^38–41^, and alcohol consumption^42–44^. This pleiotropy could suggest that these other associations are mediated by coffee intake or that these loci also influence these traits via alternative independent mechanisms. Genetic correlations have also been reported between coffee intake with other substance use^45,46^, reduced gray matter volumes^18^, psychiatric illness^45^, osteoarthritis^47^, sleep^48^, body mass index (**BMI**)^49^, type II diabetes^49^, and migraine^50^. However, some genetic correlations were conducted under *a priori* justification (e.g., other substance use traits, sleep) and as such may fail to capture the full scope of genetic correlations between coffee with other traits. Thus, a data-driven examination of trait associations with coffee intake remains unexplored.

While coffee is the primary source of caffeine for many, other common dietary sources of caffeine include tea, soft drinks, and chocolate. Consequently, when we refer to coffee intake, we mean explicit measures of coffee intake (e.g., measured as cups/day) and not caffeine intake unless otherwise specified. Intake of other caffeine sources also varies by geographic region based on beverage sales^2^. For example, tea (rather than coffee) is the preferred source of caffeine in the United Kingdom (UK; tea vs. coffee: ∼50% vs. 20%) compared to the United States (US; ∼10% vs. 30%)^2^. As some genetic studies used data from the UK Biobank (**UKB**) only^18,47,48,51–53^ or combined cohorts across regions with different patterns of caffeinated beverage intake (**Supplementary Table 1**)^32,33,46^, this distinction may limit generalizability or introduce environmental and cultural confounds that affect the genetic associations between coffee intake and other traits.

In this study, we used survey responses from US-based 23andMe, Inc. research participants of European ancestry (*N=*130,153) and performed a GWAS of a single item “How many 5-ounce (cup-sized) servings of caffeinated coffee do you consume each day?”. Using genetic correlations and phenome- and laboratory-wide association studies (**PheWAS**, **LabWAS**), we explored the relationships between coffee intake and thousands of biomarkers, health features, and lifestyle traits to provide a fuller inventory of genetic correlations with coffee intake. We compared our findings from the 23andMe cohort to those from the UKB using publicly available GWAS summary statistics of coffee intake (“How many cups of coffee do you drink each day? (Include decaffeinated coffee)”, *N=*334,659, http://www.nealelab.is/uk-biobank/). Although we had originally intended to perform a meta-analysis, our results revealed a lower-than-expected genetic correlation between coffee intake in the two cohorts; therefore, we instead used these datasets to explore cohort differences in coffee intake across these two distinct populations.

## Results

### GWAS in the 23andMe US-based cohort replicated seven loci implicated in coffee intake

Participant demographics of the 23andMe cohort are described in **Supplementary Table 2**. The cohort was 65% male, had a mean age of 52.8 ± 16.9 years old, and an average BMI of 28.38 ± 6.54 (range: 14.0-69.1), similar to the US average of 27.5 (95% CI: 25.5-29.4)^54^. The average coffee intake in the cohort was 1.98 (± 2.35 SD) cups per day, similar to the coffee intake distributions in UKB (2.14 ± 2.09 SD; see **Supplementary Figure 1** and **Supplementary Table 3** for distributions).

We conducted a GWAS of 14,274,006 imputed genetic variants assuming an additive genetic model that included age, sex, the first five genetic principal components, and indicator variables for genotype platforms as covariates (**Supplementary Table 4**; **Supplementary Material** for additional genotyping and GWAS details). The genomic control inflation factor of the GWAS was λ=1.09, suggesting no substantial inflation due to population stratification. SNP-heritability of coffee intake via Linkage Disequilibrium score regression (**LDSC**) was 7.57% ± 0.59 (**Supplementary Table 5**).

We identified seven genome-wide significant (*p<*5.00E-08) independent (*r*^2^*<*0.1) loci that were associated with coffee intake (**Figure 1**, **Table 1**; **Supplementary** Figures 2-8 for locus zoom plots). These associations replicated prior coffee or caffeine GWAS findings (**Supplementary Table 6**). For example, rs2472297 (*p=*3.60E-65, chr15q24.1) is in the intergenic region between *CYP1A1* and *CYP1A2*, and has been previously associated with coffee and caffeine intake^18,20,24,26,28,30,32,33,55^. *CYP1A1* and *CYP1A2* encode members of the cryptochrome P450 superfamily of enzymes involved in xenobiotic metabolism^22^. rs2472297 has also been previously associated with traits like alcohol consumption^56,57^, clozapine pharmacokinetics^58^, kidney function^59–62^, and the concentration of biomarkers in urine^63–68^. We also identified rs4410790 (*p=*5.20E-55, chr7p21.1), which is located upstream of the *AHR* gene encoding a transcription factor that regulates *CYP1A1/CYP1A2* and is activated by polycyclic aromatic hydrocarbons, which are present in coffee^22,69^. Prior studies associated rs4410790 and caffeine intake from tea^20^, as well as with traits like caffeine metabolism^28^, bitter beverage intake^26^, and urine biomarkers^64,66–68,70^. Lastly, rs199612805 (*p=*1.80E-10, chr22q11.23), which is located near *ADORA2A*, was also implicated in coffee intake. This variant was recently associated with caffeine intake from tea and coffee in the UKB^20^. *ADORA2A* encodes an adenosine G-protein coupled receptor that is inhibited by caffeine to produce stimulating effects^71^. The remaining four SNPs – rs34645063, rs28634426, rs117824460, and rs11474881 – were in linkage disequilibrium (**LD**) with SNPs previously identified by other coffee or caffeine GWAS^18,20,24,26,28,30,55^. rs34645063 (*p=*3.30E-09, chr6q16.1) is a deletion/insertion polymorphism between *MMS22L* and *POU3F2*. rs34645063 is in LD (R^2^=0.74) with rs754177720 and is also associated with caffeine intake from coffee or tea^20^. rs28634426 (*p=*2.10E-10, chr7q11.23) is an intronic variant of *STYXL1* in LD with rs17685 (R^2^=0.78) and rs1057868 (R^2^=0.76), which were previously implicated by coffee GWAS^18,20,24,26,30,55^. rs117824460 (*p=*1.70E-08, chr19q13.2) is an intronic variant of *CYP2A6*, and is in LD (R^2=^0.05) with rs56113850, which was implicated in coffee intake^20,26^ and caffeine metabolism^28^. *CYP2A6* encodes a cryptochrome P450 superfamily enzyme member that metabolizes nicotine^72^; rs117824460 has also been associated with smoking traits^57,73^ and serum albumin^74^, C-reactive protein^75^, and liver alkaline phosphatase levels^66^. The final significant variant we identified, rs11474881 (chr20q13.33, *p=*1.10E-08), is an intronic variant of the *PCMTD2* gene; rs11474881 is in LD (R^2^=0.98) with rs6062679, which was previously implicated in coffee and tea intake and bitter beverage consumption^26^. We used three additional multi- ancestral cohorts to replicate these findings (**Table 1**; **Supplementary Table 3**). Of the SNPs that passed QC, all replicated in a larger sample of 23andMe research participants of European ancestry (*N*=689,661), all replicated in those with African American ancestry (*N=*32,312), and one replicated in those of Latin American ancestry (*N=*124,155).

**Figure 1.**
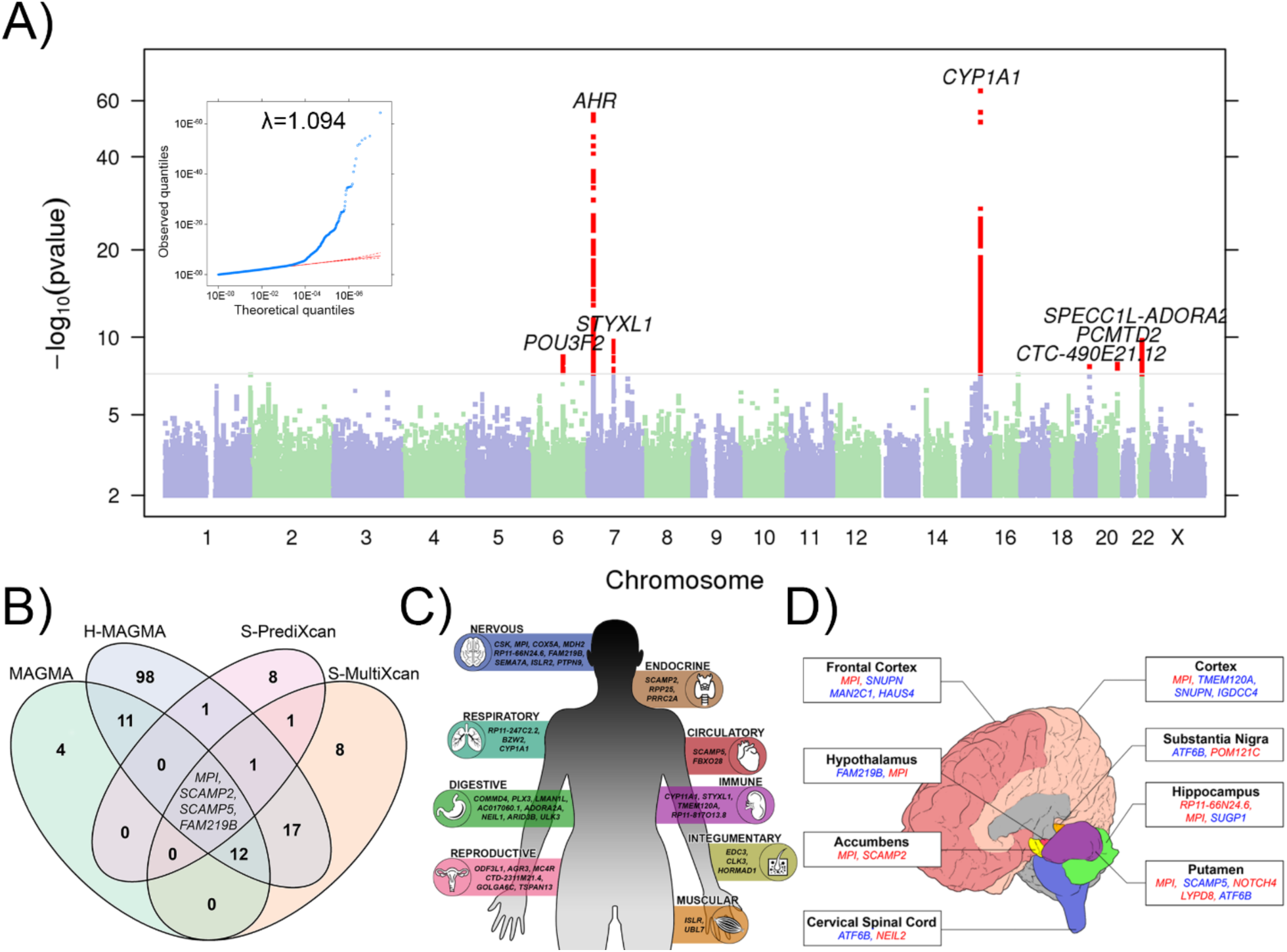
GWAS and secondary analyses of coffee intake from the 23andMe cohort. **A)** Manhattan plot displays seven genome-wide significant loci for coffee intake in the 23andMe cohort (*N=*130,153). The horizontal line represents the threshold for significance (*p=*5.00E-08). Nearest protein-coding genes (<1Mb) to significant loci are labeled. Quantile-quantile plot shown in upper left corner. For more details, see **Table 1** and **Supplementary Table 6**. **B)** Overlap of genes identified by MAGMA, H-MAGMA, S-PrediXcan, and S-MultiXcan. Genes identified by all four methods are displayed. **C)** Genes predicted to affect coffee intake identified by S-MultiXcan according to the most significantly associated biological systems. For more details, see **Supplementary Table 9. D)** Genes implicated in coffee intake by S-PrediXcan according to brain regions. Upregulated genes are shown in red, downregulated shown in blue. For more detail, see **Supplementary Table 10**.

**Table 1.** Significant (*p<*5.00E-08) GWAS results for coffee intake from 23andMe research participants (*N=*130,153) of European ancestry (**EA**). Replication (**EA Rep**) was conducted in an additional cohort of 23andMe participants of EA (*N*=689,661), and in those with African American ancestry (**AA**; *N=*32,312) and Latin American ancestry (**LA**; *N=*124,155); *SNPs that did not pass QC in replication. See **Supplementary** Table 6 for additional information.

| SNP | BP | Alleles | Cytoband | $p$ value | EA EAF | Effect | EA Rep $p$ value | EA Rep EAF | AA $p$ value | AA EAF | LA $p$ value | LA EAF | Nearest gene(s) |
| --- | --- | --- | --- | --- | --- | --- | --- | --- | --- | --- | --- | --- | --- |
| rs2472297 | 75027880 | C/T | chr15q24.1 | 3.60E-65 | 0.23 | 0.08 | 1.28E-234* | 0.24 | 3.09E-05 | 0.07 | 5.47E-47* | 0.14 | CYP1A1, CYP1A2 |
| rs4410790 | 17284577 | C/T | chr7p21.1 | 5.20E-55 | 0.38 | -0.06 | 7.58E-212* | 0.38 | 4.52E-15 | 0.52 | 9.89E-60* | 0.55 | AGR3, AHR |
| rs199612805 | 24843991 | D/I | chr22q11.23 | 1.80E-10 | 0.01 | -0.10 | 2.48E-29 | 0.015 | 1.28E-07 | 0.08 | 6.32E-13 | 0.02 | ADORA2A, UPB1 |
| rs28634426 | 75675594 | G/T | chr7q11.23 | 2.10E-10 | 0.20 | 0.03 | 3.08E-16 | 0.24 | 0.08 | 0.32 | 8.17E-06 | 0.27 | STYXL1 |
| rs34645063 | 9859107<br>5 | D/I | chr6q16.1 | 3.30E<br>-09 | 0.47 | -0.02 | 5.23E-16 | 0.48 | 0.05 | 0.6<br>5 | 4.35E-06* | 0.5<br>8 | *MMS22L,*<br>*POU3F2* |
| rs11474881 | 6289295<br>6 | D/I | chr20q13.3<br>3 | 1.10E<br>-08 | 0.55 | -0.02 | 2.36E-12 | 0.55 | .21 | 0.4<br>5 | 0.02* | 0.6<br>2 | *PCMTD2* |
| rs11782446<br>0 | 4137148<br>0 | A/G | chr19q13.2 | 1.70E<br>-08 | 0.03 | -0.06 | 2.00e-07 | 0.03 | 0.08 | 0.0<br>1 | 0.08 | 0.0<br>2 | *CTC-*<br>*490E21.1*<br>*2* |

We also report several notable nominal associations with coffee intake (*p<*1.00E-06, **Supplementary Table 6**). rs72790130 (*p=*5.50E-08, chr16q23.3) and rs2155645 (*p=*9.80E-07, chr11q23.2) are intronic variants of two cell adhesion molecule genes, *CDH13* and *NCAM1*, respectively. Both genes have been previously implicated in substance use traits by other GWAS^20,57,76–82^ and candidate gene studies in humans and animal models^83–88^. rs2155645 is also in LD with rs2298527 (R^2^=0.43), which was previously implicated in daily caffeine intake from coffee^20^. rs11204734 (*p=*2.90E-07, chr1q21.3) is an intronic variant of *ARNT*; its protein heterodimerizes with *AHR* and binds to xenobiotic response elements to regulate transcription of *CYP1A1* and *CYP1A2*^22^. Finally, rs340019 (*p=*2.10E-07, chr15q22.2) is an intronic variant of *RORA*, which is involved in circadian rhythm and metabolic regulation, among other functions^89^. rs340019 is in LD with rs12591786 (R^2^=0.25), which was implicated in daily caffeine intake from cups of coffee and tea^20^.

### Gene-based and tissue enrichment analyses suggest coffee intake is primarily associated with gene expression in the brain

We used gene- and transcriptome-based analyses (MAGMA, H-MAGMA, S-MultiXcan/S-PrediXcan) and identified 165 target candidate genes that may be most relevant to coffee intake. MAGMA identified 31 genes implicated in coffee intake in physical proximity to GWAS loci (**Supplementary Table 7**). H-MAGMA, which maps SNPs to genes via chromatin interaction from human brain tissue, implicated 143 unique gene-tissue pairs showing expression specific to cell type (75.16% neuron [31.30% cortical neuron, 33.04% iPSC derived neurons; 35.65% midbrain dopamine neurons], 24.83% astrocyte) and developmental (48.00% fetal, 52.00% adult) (**Supplementary Table 8**). Finally, S-MultiXcan predicted significant transcriptional regulation of 40 genes implicated in coffee intake dispersed across 20 tissues (**Figure 1C**; **Supplementary Table 10**). Of the top biological systems implicated by S-MultiXcan, nine were attributed to the nervous system (brain *N=*5; tibial nerve *N=*4), eight to the digestive system (esophagus *N=*6; pancreas *N=*1; small intestine *N=*1), and six to the reproductive system (testis *N=*4; prostate *N=*2; **Figure 1C**). Fifty percent of these genes were predicted to be downregulated in the digestive and reproductive systems, whereas 66.67% of nervous system genes were predicted to be upregulated. Cortical enrichment was further supported by S-PrediXcan (**Figure 1D**), showing that SNPs associated with coffee intake most frequently correlated with predicted gene expression in overall cortical and frontal cortical regions (*N=*4/tissue), as well as the putamen (*N=*5). Overall, four genes (*SCAMP2, SCAMP5, MPI,* and *FAM219B*) were identified by all four methods, and six of the 165 discovered genes (*FBXO28*, *NEIL2*, *HAUS4*, *IGDCC4*, *RP11- 298I3.5*, *RP11-298I3.5*) were not within 1Mb of SNPs identified by prior GWAS of coffee or caffeine traits (**Supplementary Table 11**; **Figure 1B**; **Table 2**). These novel genes have been associated with substance use (e.g., *HAUS4* and smoking initiation^73^), educational outcomes (e.g., *HAUS4* and educational attainment^90^), and biomarkers (e.g., *FBXO28* and mean platelet volume^91^; IGDCC4 and mean corpuscular volume^92^).

Next, we used MAGMA gene-set analysis to identify biological pathways that may be most strongly associated with coffee intake. This analysis revealed significant enrichment (*p=*4.75E- 07) in pathways related to the metabolism of xenobiotics or foreign substances (i.e., chemicals) (**Supplementary Table 12**).

MAGMA tissue-based enrichment analyses suggested that coffee intake was only significantly associated with brain tissue (**Supplementary** Figure 9A). More specifically, differential expression by coffee intake was enriched (*p<*9.25E-04) in the frontal cortex, overall cortex, cerebellum, and cerebellar hemispheres (**Supplementary** Figure 9B; **Supplementary Table 12**), consistent with the S-PrediXcan findings (**Supplementary Table 9)**.

### Genetic correlation and polygenic score analyses of coffee intake in US- and UK-based cohorts reveal discrepant associations with health and psychiatric traits, but consistent positive associations with substance use and obesity

To boost statistical power and identify novel genes associated with coffee intake, we sought to meta-analyze our data (**metaGWAS**) with those from the UKB using METAL^93^. As a preliminary step to determine the appropriateness of a meta-analysis, we examined the genetic correlation between coffee intake in the 23andMe and UKB cohorts. Surprisingly, the two datasets were only moderately correlated (*r_g_=*0.63, *p=*3.54E-43), although all top loci (*p<*5.0E-05) shared direction of effect and had similar effect strengths (**Supplementary** Figure 10). In addition, the estimated LDSC *SNP_h2_* heritability of coffee intake of our metaGWAS was slightly lower than for both the univariate GWAS (metaGWAS *SNP_h2_=*4.09% ± 0.26 vs. 23andMe *SNP_h2_=*7.57% ± 0.59 vs. UKB *SNP_h2_=*4.85% ± 0.33; **Supplementary Table 5**). We interpreted these results as an indication of cohort heterogeneity and proceeded to analyze genetic associations with coffee intake in each cohort independently.

To further understand these discrepancies, we performed a series of genetic correlation and polygenic score analyses. First, we examined the genetic architecture of coffee intake measured in 23andMe and UKB by comparing patterns of LDSC genetic correlation (*r_g_*) with 317 traits across 20 health, psychiatric, and anthropologic categories from publicly available GWAS summary statistics (**Figure 2A**; **Supplementary Table 14**). After accounting for multiple testing, 75 traits were genetically correlated with coffee intake in the 23andMe cohort and 74 traits in the UKB cohort. These associations could be underpinned by other unmeasured factors, like sugar intake from coffee sweeteners or smoking and alcohol use^95^. However, these patterns of genetic correlations persisted after conditioning on dietary sugar intake, cigarettes smoked per day, and alcohol consumption measured by the Alcohol Use Disorder Identification Test (**AUDIT**; **Supplementary Tables 11-12**; **Supplementary Tables 15-16**). Strikingly, of the traits significant in at least one cohort, only 34 (29.57%) were significant in both datasets, and only 58.82% of the traits significant in both datasets shared the same direction of correlation.

**Figure 2.**
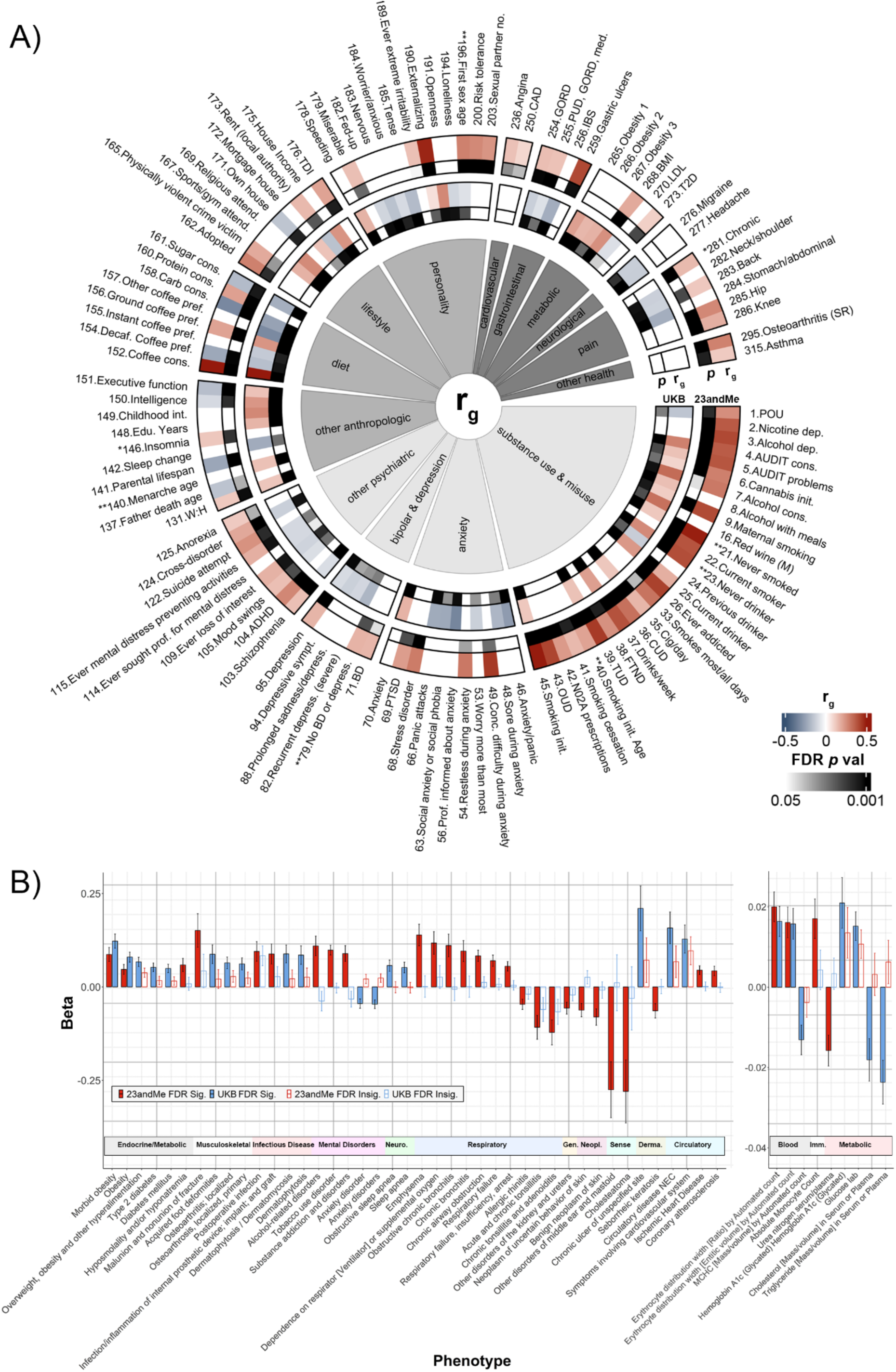
Discordant genetic and phenotypic associations with genetic disposition to coffee intake in US and UK cohorts. **A)** Comparison of genetic correlations across psychiatric (light gray), anthropologic (medium gray), and health (dark gray) traits between 23andMe (lanes 1 and 2) and UKB (lanes 3 and 4). Lanes 1 and 3 show *r_g_* values calculated by LDSC, and lanes 2 and 4 show FDR-corrected *p* values. Only traits for which at least one cohort was FDR-significant are displayed. For a full list of correlations and trait names, see **Supplementary Table 14**. Most signals persisted after conditioning for dietary sugar, cigarettes per day, and Alcohol Use Disorder Identification (**AUDIT**) Consumption scores using mtCOJO^94^ (**Supplementary Tables 15-16; Supplementary** Figures 11-12). Genetic correlations for traits denoted with * could not be calculated in both cohorts; ** denotes reverse coding. **B)** Phenomic associations (panel 1: PheWAS (*p<*3.62E-05), panel 2: LabWAS (*p<*1.57E-04)) identified from PGS of coffee intake from 23andMe and UKB summary statistics. Only traits for which at least one cohort was FDR-significant are displayed (saturated bars=FDR significant; desaturated bars=FDR non-significant). neuro.=neurological; gen.=genitourinary; neopl.=neoplasms; sense=sense organs; derma.=dermatologic; imm.=immune. For full trait names and more detail, see **Supplementary Table 18-19**.

Among the traits that were significant and consistent in direction for both cohorts, we observed positive genetic correlations between coffee intake and substance use phenotypes. For example, we identified positive genetic correlations with smoking initiation (23andMe: *r_g_=*0.50, *p=*4.74E-47; UKB: *r_g_=*0.12, *p=*1.89E-06), drinks per week (23andMe: *r_g_=*0.39, *p=*3.38E-28; UKB: *r_g_=*0.21; *p=*1.39E-14), and cannabis initiation (23andMe: *r_g_=*0.28, *p=*1.34E-08; UKB: *r_g_=*0.09, *p=*5.61E-03). The strength of genetic correlations for substance use and misuse traits significant in at least one cohort was stronger in 23andMe compared to the UKB (0.30±0.03 vs. 0.09±0.02; Welch’s *t*(51.97)=5.96, *p=*2.23E-07). For example, associations with substance use disorder and dependence traits were mostly observed in the 23andMe cohort and were weaker or not observed in the UKB, such as for tobacco use disorder, opioid use disorder, cannabis use disorder, nicotine dependence, and alcohol dependence (*r_g_=*0.24 to 0.44, *p=*6.54E-23 to 2.12E-03), as well as externalizing (23andMe: *r_g_=*0.48, *p=*7.21E-41; UKB: *r_g_=*0.07, *p=*4.37E-03), which is highly correlated with substance use and misuse^96^. Cluster analysis showed that genetic correlations for coffee intake in both cohorts aligned more with general substance use than misuse (**Supplementary** Figure 13).

Metabolic traits were largely congruent in their positive genetic correlations with coffee intake in both cohorts. For example, BMI (23andMe: *r_g_=*0.19, *p=*1.61E-11; UKB: *r_g_=*0.25, *p=*7.85E- 26) and waist-to-hip ratio (23andMe: *r_g_=*0.12, *p=*4.33E-04; UKB: *r_g_=*0.13, *p=*3.96E-07) were positively genetically correlated with coffee intake in both datasets. Also consistent across cohorts were the lack of significant genetic correlations with most cardiovascular and cancer traits.

The majority of traits were only significant in one cohort or showed discrepancies in the direction of association. For example, coffee intake measured in the 23andMe dataset was positively genetically correlated with anxiety-related traits (*r_g_=*0.22 to 0.44, *p=*1.41E-05 to 8.53E- 03). In contrast, all significant genetic correlations between coffee intake and anxiety-related traits in the UKB were negative (*r_g_*= -0.33 to -0.12, *p=*5.49E-06 to 8.12E-03), except clinically diagnosed anxiety (*r_g_=*0.17, *p=*1.39E-05). We also identified significant positive genetic correlations with cross-disorder, attention deficit hyperactivity disorder, schizophrenia, and anorexia (*r_g_=*0.12 to 0.27, *p=*1.00E-07 to 0.01) that were exclusive to the 23andMe dataset, whereas these associations were not apparent or were negatively genetically correlated in the UKB (*r_g_=* -0.13 to 0.02, *p=*1.01E-04 to 0.55). Significant positive correlations with cognitive variables, such as executive function and intelligence, were found in the UKB (*r_g_=* 0.13 to 0.23, *p=*4.55E-08 to 8.04E- 23), though these were negatively genetically correlated in 23andMe (*r_g_=* -0.17 to -0.10, *p=*7.83E- 08 to 2.06E-03). Certain correlations with physical health traits also differed between cohorts. While correlations with most gastrointestinal traits in the 23andMe cohort were positive, such as a positive genetic correlation with gastric ulcers (*r_g_=*0.41, *p=*3.58E-03), the corresponding genetic correlations observed in the UKB were either non-significant or negative (*r_g_*= -0.22 to 0.12, *p=*1.34E-06 to 0.88). Positive genetic correlations with chronic pain as well as back, hip, and knee pain were observed in the 23andMe dataset (*r_g_=*0.12 - 0.26, *p=*9.02E-08 - 3.58E-03), yet only negative genetic correlations with pain traits were reported in the UKB (*r_g_=*-0.22 to -0.12, *p=*6.23E- 04 - 2.54E-06). Across all health and psychiatric traits that were significant within each cohort, all traits showed a positive genetic correlation with coffee intake in 23andMe participants. Only 41.3% of correlations were positive in the UKB.

We observed similar discrepancies when we extended our results to a health-care system population (**Figure 2B**). We conducted PheWAS and LabWAS by testing the association between polygenic scores (**PGS**) for coffee intake derived from 23andMe or the UKB with 1,655 medical traits and biomarkers. We identified 31 PheWAS and LabWAS traits that met the 5% FDR significance threshold using the 23andMe PGS, and 24 using the UKB PGS (**Supplementary Tables 17 and 18**). Only two endocrine traits (i.e., obesity and morbid obesity) and two biomarkers related to red blood cells were consistent in significance and direction of association. Otherwise, all significant associations were observed when testing PGS generated from one cohort but not the other. For instance, when coffee intake PGS were derived from 23andMe, among the top positive PheWAS and LabWAS associations were substance use disorders and respiratory conditions (e.g., chronic airway obstruction, emphysema, and respiratory failure) and absolute monocyte count. Among the top negative associations derived from 23andMe PGS were those with sense organs, neoplasms, certain respiratory conditions (i.e., allergic rhinitis, acute and chronic tonsillitis, chronic tonsillitis and adenoiditis), and urea nitrogen serum/plasma. When coffee intake PGS were derived from UKB, among the top positive PheWAS and LabWAS associations were endocrine and musculoskeletal disorders, as well as the two metabolic biomarkers, glycated hemoglobin A1c and glucose. The only significant negative PheWAS and LabWAS associations from UKB-derived PGS were with anxiety disorders, and biomarkers related to blood (mean corpuscular hemoglobin concentration) and metabolic (cholesterol and triglycerides in serum or blood) traits.

## Discussion

In this study, we contributed to the existing GWAS literature of coffee intake by analyzing a US population of 130,153 participants. We uncovered seven loci associated with coffee intake, most of which were in genes implicated in metabolic processes. Coffee related variants were significantly enriched in the central nervous system. Despite prior evidence that coffee intake confers health benefits, we found genetic correlations mostly with adverse outcomes in both cohorts, particularly substance use disorders and obesity-related traits, in both cohorts. Relationships with other medical, anthropologic, and psychiatric traits were inconsistent in the US and UK cohorts, suggesting that differences between populations may affect coffee intake GWAS results and its genetic relationships with other traits.

Our GWAS replicated prior associations with genes and variants implicated in coffee and caffeine intake as well as other metabolic and xenobiotic processes^28^, including rs2472297 near *CYP1A1/CYP1A2*^18,24,26,33,46^ and rs4410790 near *AHR*^18,23,24,26,27,46,97^, even though our study sample was smaller compared to other GWAS (*N=*125,776-373,522^20,24,26^). Gene-based analyses uncovered 165 candidate genes, including four genes that overlapped across all four analyses: *MPI, SCAMP2, SCAMP5*, and *FAM219B*, all of which have been implicated in a prior coffee GWAS^18^. These overlapping genes have other associations with substance use and medical biomarkers including blood pressure, hypertension, and LDL cholesterol^91,98–104^. We identified gene enrichment in brain tissues across the frontal cortex, putamen, and hippocampus, consistent with prior GWAS showing enrichment for SNPs associated with coffee and caffeine in the central nervous system^18,20,26^. This is supported by brain imaging studies across cortical and subcortical areas showing morphological^105–108^ and functional^109,110^ differences between those who habitually drink coffee compared to those who do not.

One of the most striking observations of this study is the breadth and magnitude of positive associations between coffee intake with substance use. It is widely believed that use of one substance heightens risk for use of other substances and that there are common genetic risk factors for any substance use^111,112^; coffee, which is not generally considered a drug of misuse, does not appear to be exempt from this. We identified positive genetic correlations between coffee intake and other substances (i.e., tobacco, alcohol, cannabis and opioid use), as well as relevant personality traits like externalizing behavior. The genetics of coffee intake aligned with substance consumption phenotypes, corroborating prior GWAS and twin studies^113–115^ (but see^23^), but not with substance misuse. This is perhaps unsurprising because the phenotype probed by the 23andMe and UKB cohorts focuses on quantity rather than clinically-defined dependence. We and others previously demonstrated that the genetic architectures of other substance intake versus problematic use are unique^43,111,116–119^, and this is likely also true for coffee.

We found consistent positive genetic correlations with BMI and obesity in both 23andMe and the UKB. This is in contrast to meta-analyses of randomized control trials and epidemiological studies that found unclear effects by any coffee or decaffeinated coffee intake on waist circumference and BMI-defined obesity, and a modest inverse relationship between coffee intake and BMI^120,121^. Results for these studies are highly heterogeneous, likely due to interindividual variability in the inclusion of sugary coffee additives, cultivation, roasting, and brewing conditions affecting its chemical makeup^9,122^, and other habits surrounding coffee intake (e.g., concurrent food intake or appetite suppression by nicotine if smoking concurrently^123^). This contentious relationship may also be explained by the amount of coffee intake, as greater coffee intake seems to attenuate the genetic associations with BMI and obesity^49^, possibly due to the appetite suppressant effects of caffeine^124^. Alongside accounting for other dietary intake, detailed accounting of coffee preparation, and consumptive habits formed with coffee intake, future subgroup analyses may help explain discrepant associations between the genetics and prevalence of coffee intake with BMI-related traits.

We did not recapitulate the beneficial phenotypic relationships between coffee intake and a variety of health outcomes that are generally reported by health association studies^6–8,10,125–137^, perhaps because our study focused on the genetic relationship between coffee intake and other medical outcomes, or because our study focused on coffee intake and not caffeine intake. At the genetic level, we find no evidence of a common genetic background that could explain the beneficial effects of coffee on 29 cancers, Alzheimer’s disease/dementia/cognitive impairments, Parkinson’s disease, diabetes, cirrhosis, most cardiovascular conditions, or gout. In fact, some of these associations (e.g., cardiovascular traits and type II diabetes) were positive in the 23andMe cohort but showed no significant associations in the UKB cohort. Similarly, phenome-wide analysis did not support prior cancerous, metabolic, cardiovascular, or neurological health advantages of coffee intake^6–8,10,126–136,138^. Although this may seem discrepant to phenotypic associations that generally report health benefits of coffee intake, recent meta-analysis of over 100 phenotypic studies on coffee intake health outcomes suggest high levels of heterogeneity across cohorts^6,7^, especially across geographically separated populations^6^.

We found many opposing relationships with the genetics of coffee intake between 23andMe and UKB. For example, genetic correlations with pain, psychiatric illnesses, and gastrointestinal traits were positively genetically correlated with coffee intake in 23andMe, but these associations were negative in the UKB. Inversely, the UKB analysis revealed that coffee intake was positively genetically correlated with cognitive traits, such as executive function and intelligence, corroborating prior evidence^139–142^, yet genetic correlations with these two traits were negative in 23andMe. Multiple PheWAS associations were also discordant. When PGS were derived from 23andMe, we observed heightened odds between genetic liability for coffee intake and respiratory illnesses, ischemic heart disease, infection, and alcohol-related disorders. Higher odds for musculoskeletal and sleep conditions were mostly associated with coffee PGS generated from the UKB. Only 11 out of the 42 phenotypes associated with coffee intake PGS showed negative associations, and none of these purported health “benefits” were consistently observed in both cohorts. Whereas the coffee intake PGS from 23andMe was associated with lower odds for ear conditions, skin neoplasms, allergic rhinitis, and tonsillitis, the PGS of coffee intake from the UKB was associated with a lower risk of anxiety disorders. Also of note is that the number of positive genetic correlations and PGS associations between coffee intake and these other traits was greater when analyzed using data from the 23andMe cohort versus from the UKB, and the strength of these associations was usually stronger. Partially consistent with this, one meta-analysis of mortality found an inverse relationship between coffee intake and all-cause mortality in European but not US studies^143^.

Our study shows that cultural, cohort, or geographic influences could affect the inferred genetic architecture of coffee intake and its associations with other health and lifestyle outcomes. Geographic regions may have an observable influence on GWAS results^144^. We observed no significant differences in subtle geographic differences on coffee intake correlations using location data available in the UKB (**Supplementary** Figure 14), suggesting cultural differences may contribute more to the cohort variations we report here. There is considerable variation in how or with whom one may consume coffee that could be subject to cultural influence. Caffeinated beverage sales, for instance, suggest that coffee and carbonated caffeinated beverages are more preferred in the US than the UK^2^, whereas tea is the preferred source of caffeine in the UK and may modify coffee intake^2^ (**Supplementary** Figure 15). Higher levels of coffee intake or caffeine from high caloric beverages in the US cohort may partially explain the higher number and magnitude of negative health associations observed in the 23andMe analysis. Even across coffee beverage subtypes, the concentration of caffeine, other coffee chemical constituents, and manufacturing byproducts (e.g., plastics and metals from packaging) varies and thus may be important parameters in health associations^122,145,146^. A recent investigation revealed the volume of ground or instant coffee is important to the potential health effects of its intake^147^; instant coffee (∼60 mg of caffeine per cup) is more commonplace in the UK whereas fresh brewed coffee (∼85 mg of caffeine per cup ^20^) is preferred in the US^2^. Cultural differences in coffee intake could help explain the divergent patterns of health and lifestyle associations between UK and US participants, though the relative contributions of culture, geography, and their interactions to these differences will need further exploration.

There are multiple caveats to consider when interpreting our findings. Firstly, our study does not address causality between coffee intake and other health and lifestyle traits. Mendelian randomization (**MR**) studies have attempted to address the exposure-outcome relationships between two traits by using genetic instruments (i.e., SNPs identified by GWAS) as proxies for exposure and associating them with an outcome of interest. For example, MR using genetic markers associated with coffee intake suggest that coffee consumption has no causal effect on obesity and endocrine disorders, despite observational studies suggesting protective effects of coffee^148^. Similarly, MR studies of coffee and other substance use (e.g., tobacco, alcohol, cannabis) are also contentious^149,150^, with evidence that inconsistencies may be driven by gene-cohort confounds such as those we found in this study^151^. Secondly, the phenotype examined by 23andMe was exclusively caffeinated coffee intake, with one cup defined as 5 ounces, whereas the UKB also included decaffeinated coffee and did not explicitly define the volume of one cup. The caffeine content within coffee was also not directly measured. However, secondary analysis using summary statistics of estimated caffeine intake from any coffee subtype in the UKB^20^ yielded remarkably similar patterns of genetic correlations as those derived from our GWAS of cups of coffee consumed (**Supplementary** Figure 16, **Supplemental Table 14**). This analysis presumably mitigated the relative contribution of decaffeinated coffee (3mg of caffeine per cup versus 60 to 85mg per cup of caffeinated coffee^20^) to the revealed genetic associations, so we do not believe the cohort discrepancies are driven by the inclusion of decaffeinated coffee drinkers in the UKB. Another consideration is the possible health effects of non-caffeine coffee components, which are comparatively under-investigated^9^, such as other coffee bean phytochemical and drink additives. Furthermore, while it is unlikely that the discrepancies in genetic associations are driven by age, which is similar between cohorts (approximately 53 years old in 23andMe versus 57^20^ years old in UKB), these cohorts skew older than the population average. They are also of above average socioeconomic status^152^ and are of European descent, limiting generalizability of our findings to a larger population. Some studies also show sex-dependent differences in coffee and caffeine metabolism and health associations with intake^138,153,154^, which was not examined in our study.

Overall, we present striking differences in genetic associations of coffee intake across two large cohorts of European ancestry. While some genetic signals replicate across diverse cohorts, such as our GWAS findings and the associations between coffee intake with substance use and obesity traits, other associations may be obscured by cohort or cultural differences related to the phenotype in question. Our study provides a cautionary perspective on combining large cohort datasets gathered from unique geo-cultural populations.

## Acknowledgements

We would like to thank the research participants and employees of 23andMe for making this work possible. The following members of the 23andMe Research Team contributed to this study: Stella Aslibekyan, Adam Auton, Elizabeth Babalola, Robert K. Bell, Jessica Bielenberg, Katarzyna Bryc, Emily Bullis, Daniella Coker, Gabriel Cuellar Partida, Devika Dhamija, Sayantan Das, Teresa Filshtein, Kipper Fletez-Brant, Will Freyman, Karl Heilbron, Pooja M. Gandhi, Karl Heilbron, Barry Hicks, David A. Hinds, Ethan M. Jewett, Yunxuan Jiang, Katelyn Kukar, Keng-Han Lin, Maya Lowe, Jey C. McCreight, Matthew H. McIntyre, Steven J. Micheletti, Meghan E. Moreno, Joanna L. Mountain, Priyanka Nandakumar, Elizabeth S. Noblin, Jared O’Connell, Aaron A. Petrakovitz, G. David Poznik, Morgan Schumacher, Anjali J. Shastri, Janie F. Shelton, Jingchunzi Shi, Suyash Shringarpure, Vinh Tran, Joyce Y. Tung, Xin Wang, Wei Wang, Catherine H. Weldon, Peter Wilton, Alejandro Hernandez, Corinna Wong, Christophe Toukam Tchakouté.

We would also like to thank The Externalizing Consortium for sharing the GWAS summary statistics of externalizing. The Externalizing Consortium: Principal Investigators: Danielle M. Dick, Philipp Koellinger, K. Paige Harden, Abraham A. Palmer. Lead Analysts: Richard Karlsson Linnér, Travis T. Mallard, Peter B. Barr, Sandra Sanchez-Roige. Significant Contributors: Irwin D. Waldman. The Externalizing Consortium has been supported by the National Institute on Alcohol Abuse and Alcoholism (R01AA015416 -administrative supplement), and the National Institute on Drug Abuse (R01DA050721). Additional funding for investigator effort has been provided by K02AA018755, U10AA008401, P50AA022537, as well as a European Research Council Consolidator Grant (647648 EdGe to Koellinger). The content is solely the responsibility of the authors and does not necessarily represent the official views of the above funding bodies. The Externalizing Consortium would like to thank the following groups for making the research possible: 23andMe, Add Health, Vanderbilt University Medical Center’s BioVU, Collaborative Study on the Genetics of Alcoholism (COGA), the Psychiatric Genomics Consortium’s Substance Use Disorders working group, UK10K Consortium, UK Biobank, and Philadelphia Neurodevelopmental Cohort.

MVJ, SBB, and SSR are supported by funds from the California Tobacco-Related Disease Research Program (TRDRP; Grant Number T29KT0526 and T32IR5226). SBB and AAP were also supported by P50DA037844. BP, JJM, and SSR are supported by NIH/NIDA DP1DA054394. HHAT is funded through a Natural Science and Engineering Research Council PGS-D scholarship and Canadian Institutes of Health Research (CIHR) Fellowship. JYK is supported by a CIHR Canada Research Chair in Translational Neuropsychopharmacology. LKD is supported by R01 MH113362. NSC is funded through an Interdisciplinary Research Fellowship in NeuroAIDs (Grant Number R25MH081482). JM is funded by the National Institute for Health and Care Research (NIHR) Maudsley Biomedical Research Centre at South London and Maudsley NHS Foundation Trust and King’s College London. The content is solely the responsibility of the authors and does not necessarily represent the official views of the National Institutes of Health.

The datasets used for the PheWAS and LabWAS analyses described were obtained from Vanderbilt University Medical Center’s BioVU which is supported by numerous sources: institutional funding, private agencies, and federal grants. These include the NIH funded Shared Instrumentation Grant S10RR025141; and CTSA grants UL1TR002243, UL1TR000445, and UL1RR024975. Genomic data are also supported by investigator-led projects that include U01HG004798, R01NS032830, RC2GM092618, P50GM115305, U01HG006378, U19HL065962, R01HD074711; and additional funding sources listed at https://victr.vumc.org/biovu-funding/. PheWAS and LabWAS analyses used CTSA (SD, Vanderbilt Resouces). This project was supported by the National Center for Research Resources, Grant UL1 RR024975-01, and is now at the National Center for Advancing Translational Sciences, Grant 2 UL1 TR000445-06.

## Author Contributions

SSR and AAP conceived the idea. PF and SLE contributed formal analyses and curation of 23andMe data. HHAT contributed to formal analyses, investigation, and data visualization. BP, AA, and NCK contributed to formal data analysis and data visualization. JJM and LVR contributed to formal analyses. JM contributed to data visualization. HHAT and SSR wrote the manuscript. All authors reviewed and edited the manuscript.

## Materials and Correspondence

Correspondence and material requests should be directed to Sandra Sanchez-Roige.

## Data availability

We will provide 23andMe summary statistics for the top 10,000 SNPs upon publication. Full GWAS summary statistics will be made available through 23andMe to qualified researchers under an agreement with 23andMe that protects the privacy of the 23andMe participants. Please visit (https://research.23andme.com/collaborate/#dataset-access/) for more information and to apply to access the data.

## Methods

### Study cohorts, coffee intake and univariate GWAS

#### 23andMe

Univariate GWAS was conducted in a sample of 130,153 male and female research participants of the genetics testing company 23andMe, Inc, as previously described^155^. Participants provided informed consent and volunteered to participate in the research online, under a protocol approved by the external AAHRPP-accredited IRB, Ethical & Independent (E&I) Review Services. As of 2022, E&I Review Services is part of Salus IRB (https://www.versiticlinicaltrials.org/salusirb). During 4 months in 2015 and 14 months between 2018-2020, participant responses to the question “How many 5-ounce (cup-sized) servings of caffeinated coffee do you consume each day?” were collected as part of a larger survey. Participants categorized as of European descent by genotype data were included in the univariate GWAS (see **Supplementary Material**)^156^. Participant demographics are presented in **Supplementary Table 2**.

DNA extraction and genotyping were performed from saliva samples by clinical laboratories CLIA-certified and CAP-accredited by the Laboratory Corporation of America. 23andMe, Inc. conducted all quality control, imputation, and univariate genome-wide analyses as previously described (see **Supplementary Table 4** for SNPs analyzed following quality control and imputation)^157,158^. Variants were imputed based on an imputation panel combining 1000 Genomes Phase 3, UK10K and the Human Reference Consortium. The 23andMe pipeline removes variants of low genotyping quality (failed a Mendelian transmission test in trios (*p*<1.00E-20), failed Hardy-Weinberg test (*p*<1.02E-20), failed batch effects test (ANOVA *p*<1.00E-20), or had a call rate <90%) or imputation quality (r^2^<0.50 averages across genotyping arrays or a minimum r^2^<0.3 or variants with apparent batch effects (*p*<1.00E-50))^157,159^. The 23andMe GWAS pipeline performs linear regression and assumes an additive model for allelic effects^43,155,160,161^. Only unrelated participants were included in the GWAS, which was determined using a segmental identity-by-descent (**IBD**) estimation algorithm. Two individuals that shared more than 700 cM IBD at one or both genomic segments (∼20% of the genome) were classified as related. This is the minimum threshold expected to identify first cousins in an outbred population. Age (inverse-normal transformed), sex, the top five principal genotype components, and indicator variables for genotype platforms were applied as covariates and *p*-values were corrected for genomic control.

We conducted replication in three multi-ancestral cohorts (European ancestry *N*=689,661; African American ancestry *N=*32,312; Latin American ancestry *N=*124,155; daily mg of caffeine from coffee, transformed by log10(x+75)). Demographic information on these cohorts is shown in **Supplementary Table 3**. Ancestry was determined by analyzing local ancestry (see **Supplementary Material**)^156^.

#### UK Biobank

Summary statistics of coffee intake (*N=*334,659) were generated from UK Biobank (**UKB**) participants. Participants provided informed consent, were of White British descent, and answered the question “How many cups of coffee do you drink each day? (Include decaffeinated coffee)”. Other previously published GWAS of coffee intake with publicly available summary statistics were not included in our meta-analysis due to differences in the way that coffee intake was measured (e.g., “How often do you drink coffee?”, “How much coffee do you consume per year?”^30^), or differences in ascertainment (e.g., Parkinson’s disease only^31^). Secondary analysis was also conducted with GWAS summary statistics of caffeine intake from coffee (*N=*373,522) in the UKB that was calculated based on the number of cups of caffeinated and decaffeinated coffee consumed^20^. For further information about the UKB data collection and GWAS summary statistics for coffee intake and caffeine consumed from coffee, see http://www.nealelab.is/uk-biobank/ (field 1498, both sexes) and Said et al. 2020^20^, respectively.

#### GWAS meta-analysis

We performed sample size weighted meta-analysis of the 23andMe and UKB cohorts using METAL (version 2020-05-05)^93^ as previously described^43^. A total of 491,347 participants of European ancestry and 9,551,852 SNPs passing quality control were included in this meta-analysis.

### Gene-based analyses (MAGMA, H-MAGMA, SPrediXcan/S-MultiXcan)

The web-based platform Functional Mapping and Annotation of Genome-Wide Association Studies (**FUMA** v1.3.8) was used to further explore the functional consequences of lead SNPs and identify prior associations in the literature. GWA significant lead SNPs of coffee cups per day consumed from the UKB were identified using FUMA. SNPs were annotated based on ANNOVAR categories, Combined Annotation Dependent Depletion scores, RegulomeDB scores, expression quantitative trait loci (**eQTLs**), and chromatin state predicted by ChromHMM. Novel coffee intake candidate genes were identified as genes not in linkage disequilibrium or within 1Mb of GWAS-significant SNPs uncovered by other GWAS of coffee and caffeine traits (e.g., coffee/caffeine intake or caffeine metabolism). These SNPs were identified using the EBI GWAS Catalog (https://www.ebi.ac.uk/gwas/).

#### MAGMA gene-based and pathway analyses

We used “Multi-marker Analysis of GenoMic Annotation” (MAGMA, v1.08) to conduct gene-based associations on the 23andMe GWAS summary statistics of coffee intake. SNPs were annotated to protein-coding genes using FUMA and Ensembl build v92, which accounts for SNP LD using multiple regression methods. The default settings were used, and LD was estimated using the 1000 Genomes European reference sample. The significance of associations across 19,773 genes were adjusted using Bonferroni correction for multiple testing (one-sided *p<*2.53E-06; **Supplementary Table 7**). Gene-set analysis was subsequently conducted on 10,678 gene-sets and Gene Ontology terms curated from the Molecular Signatures Database (MsigDB v7.0). Tissue-specific gene expression profiles were also assessed in 54 tissue types and 30 general tissue types across the body with average gene expression in each tissue type used as a covariate in the analysis (**Supplementary Table 13**). Using Genome-Tissue Expression (**GTEx**, v8) RNA-seq data, gene expression values were log_2_ transformed values of the average Reads Per Kilobase Million (RPKM) for each tissue type (RPKM>50 were listed as 50). Significance was determined following Bonferroni correction (one-sided *p<*9.26E-04 for 54 tissue types; one-sided *p<*1.67E-03 for 30 general tissue types).

#### H-MAGMA

To identify neurobiologically relevant target genes, we incorporated coffee intake GWAS data with chromatin interaction profiles from human brain tissue using Hi-C coupled MAGMA (**H-MAGMA**)^162^.

#### S-PrediXcan and S-MultiXcan

We performed a transcriptome-wide association study (**TWAS**) using the MetaXcan package (ver0.7.5)^163,164^ consisting of S-PrediXcan and S-MultiXcan to identify specific eQTL-linked genes associated with coffee intake. eQTLs are genomic loci that contribute to heritable variation in mRNA levels that might influence the expression of a particular gene or its neighbors. This approach uses genetic information to predict gene expression levels in various brain tissues and tests whether the predicted gene expression correlates with coffee intake. S-PrediXcan uses precomputed tissue weights from the GTEx project database (https://www.gtexportal.org/) as the reference transcriptome dataset via Elastic net models. As input data, we included our GWAS summary statistics, transcriptome tissue data, and covariance matrices of the SNPs within each gene model (based on HapMap SNP set; available to download at the PredictDB Data Repository) from all available tissues (*N=*49). We applied a Bonferroni correction for multiple testing across all tissues (*N=*21,565; **Supplementary Table 10**).

### LDSC heritability and genetic correlations

Linkage Disequilibrium Score regression (**LDSC**; https://github.com/bulik/ldsc) was used to calculate heritability (***SNP-h_2_***) and genetic correlations between habitual coffee intake and other phenotypes^165^. *SNP-h_2_* was calculated from publicly available, pre-computed LD scores (“eur_w_ld_chr/”). LDSC was also used to calculate genetic correlations (***r_g_***) between habitual coffee intake and 317 selected traits informed by prior literature across the following categories: substance use and misuse, anxiety, bipolar disorder & depression, cancer, cardiovascular, diet, gastrointestinal, lifestyle, metabolic, neurological, pain, personality, other anthropologic, other health, and other psychiatric traits.

### mtCOJO

We conditioned summary statistics on traits that are correlated with coffee intake using mtCOJO^94^ to determine if genetic associations are retained after controlling for their effects. Conditioning on GWAS summary statistics for dietary sugar intake^166^, Alcohol Use Disorder Identification Test (**AUDIT**) consumption^43^, and cigarettes smoked per day^73^ were examined at a *p<*1.0E-5 and clump-r^2^ threshold of 0.10.

### Phenome and laboratory-wide association studies

We conducted phenome-wide association analyses (**PheWAS**) and Laboratory-wide association analyses (**LabWAS**) to test the association between polygenic scores (**PGS**) for coffee intake and liability across thousands of medical conditions from hospital-based cohorts. These analyses were conducted using data from the Vanderbilt University Medical Center (**VUMC**). The project was approved by the VUMC Institutional Review Board (IRB #160302, #172020, #190418). VUMC is an integrated health system with individual-level health data from electronic health record (**EHR**) data for about 3.2 million patients. The VUMC biobank contains clinical data from EHR as well as biomarkers obtained from laboratory assessments. A portion of the individuals from VUMC also have accompanying array genotyping data. This cohort, with over 72,821 patients, is called BioVU^167,168^.

For each of the unrelated genotyped individuals of European ancestry from BioVU, we computed polygenic scores for coffee intake using the PRS-CS “auto” version^167^. Genotyping and quality control for this cohort have been extensively described^168,169^.

#### Phenome-wide association analyses (PheWAS)

To identify associations between the PGS for coffee and clinical phenotypes, we performed a PheWAS. We fitted a logistic regression model to each of 1,338 case/control disease phenotypes (“phecodes”) to estimate the odds of each diagnosis given the coffee PGS, while adjusting for sex, median age of the longitudinal EHR, and the first 10 PCs. Analyses were conducted using the PheWAS v0.12 R package^170^. We required the presence of at least two International Disease Classification codes mapped to a PheWAS disease category (Phecode Map 1.2; https://phewascatalog.org/phecodes) and a minimum of 100 cases for inclusion of a phecode. The disease phenotypes included 145 circulatory system, 120 genitourinary, 119 endocrine/metabolic, 125 digestive, 117 neoplasms, 91 musculoskeletal, 85 sense organs, 76 injuries & poisonings, 65 dermatological, 76 respiratory, 69 neurological, 64 mental disorders, 42 infectious diseases, 42 hematopoietic, 34 congenital anomalies, 37 symptoms, and 31 pregnancy complications.

#### Laboratory-wide association analyses (LabWAS)

We also examined laboratory results in BioVU, which we refer to as LabWAS. We implemented the pipeline already established by Dennis, et al. ^169^. Broadly, LabWAS uses the median, inverse normal quantile transformed age-adjusted values from the QualityLab pipeline in a linear regression to determine the association with the input coffee intake PGS variable. We controlled for the same covariates as for the PheWAS analyses, excluding median age because the pipeline corrects for age using cubic splines with 4 knots.

### Cluster analysis

Previous studies have shown that consumption and misuse/dependence phenotypes have a distinct genetic architecture^43,111,116–119^. To explore whether the coffee intake analysis clustered closer to substance intake or misuse/dependence phenotypes, we used an unsupervised machine learning hierarchical clustering algorithm known as agglomerative nesting (**AGNES**)^167^ on a genetic correlation matrix of all traits. AGNES initially forms single-item clusters that are fused together into intermediate groups until all traits are included in a single cluster^171^. Clusters are formed with Ward’s method such that the total within cluster variance is minimized while maintaining the fewest number of clusters based on cluster dissimilarity. Dissimilarity is assessed through Euclidean Distance of each pairwise genetic correlation with another trait. The product of AGNES is a dendrogram, formed with multiple brackets called “branches”. AGNES was implemented in R using the *cluster* package (ver2.1.4)^167^.

Clustering was conducted with summary statistics of cigarettes per day^73^, former smoker^73^, smoking initiation^73^, problematic opioid use^161^, ICD10 F17 nicotine dependence^172^, alcohol dependence^173^, AUDIT consumption^116^, AUDIT problems^116^, cannabis initiation^79^, cannabis use disorder^118^, drinks per week^73^, externalizing psychopathology^96^, Fagerström Test for Nicotine Dependence (FTND)^174^, general risk tolerance^175^, age of smoking initiation^73^, and opioid use disorder^161^. The genetic correlations of cigarettes per day, former smoker, and smoking initiation were reverse coded to show the intuitive effects against the other traits in the dendrogram.

## References

1 International Coffee Organization. Annual Review Coffee Year 2019/2020. (International Coffee Organization, London, 2021).

2 Reyes, C. M. & Cornelis, M. C. Caffeine in the Diet: Country-Level Consumption and Guidelines. Nutrients 10, doi:10.3390/nu10111772 (2018).

3 Landais, E. et al. Coffee and Tea Consumption and the Contribution of Their Added Ingredients to Total Energy and Nutrient Intakes in 10 European Countries: Benchmark Data from the Late 1990s. Nutrients 10, doi:10.3390/nu10060725 (2018).

4 Rehm, C. D., Ratliff, J. C., Riedt, C. S. & Drewnowski, A. Coffee Consumption among Adults in the United States by Demographic Variables and Purchase Location: Analyses of NHANES 2011-2016 Data. Nutrients 12, doi:10.3390/nu12082463 (2020).

5 Camandola, S., Plick, N. & Mattson, M. P. Impact of Coffee and Cacao Purine Metabolites on Neuroplasticity and Neurodegenerative Disease. Neurochem Res 44, 214–227, doi:10.1007/s11064-018-2492-0 (2019).

6 Grosso, G., Godos, J., Galvano, F. & Giovannucci, E. L. Coffee, Caffeine, and Health Outcomes: An Umbrella Review. Annu Rev Nutr 37, 131–156, doi:10.1146/annurev-nutr-071816-064941 (2017).

7 Poole, R. et al. Coffee consumption and health: umbrella review of meta-analyses of multiple health outcomes. BMJ 359, j5024, doi:10.1136/bmj.j5024 (2017).

8 Kositamongkol, C. et al. Coffee Consumption and Non-alcoholic Fatty Liver Disease: An Umbrella Review and a Systematic Review and Meta-analysis. Front Pharmacol 12, 786596, doi:10.3389/fphar.2021.786596 (2021).

9 Socala, K., Szopa, A., Serefko, A., Poleszak, E. & Wlaz, P. Neuroprotective Effects of Coffee Bioactive Compounds: A Review. Int J Mol Sci 22, doi:10.3390/ijms22010107 (2020).

10 Zhao, L. G. et al. Coffee drinking and cancer risk: an umbrella review of meta-analyses of observational studies. BMC Cancer 20, 101, doi:10.1186/s12885-020-6561-9 (2020).

11 Freedman, N. D., Park, Y., Abnet, C. C., Hollenbeck, A. R. & Sinha, R. Association of coffee drinking with total and cause-specific mortality. N Engl J Med 366, 1891–1904, doi:10.1056/NEJMoa1112010 (2012).

12 Svikis, D. S. et al. Coffee and energy drink use patterns in college freshmen: associations with adverse health behaviors and risk factors. BMC Public Health 22, 594, doi:10.1186/s12889-022-13012-3 (2022).

13 Swan, G. E., Carmelli, D. & Cardon, L. R. Heavy consumption of cigarettes, alcohol and coffee in male twins. J Stud Alcohol 58, 182–190, doi:10.15288/jsa.1997.58.182 (1997).

14 Treur, J. L. et al. Associations between smoking and caffeine consumption in two European cohorts. Addiction 111, 1059–1068, doi:10.1111/add.13298 (2016).

15 Tang, N., Wu, Y., Ma, J., Wang, B. & Yu, R. Coffee consumption and risk of lung cancer: a meta-analysis. Lung Cancer 67, 17–22, doi:10.1016/j.lungcan.2009.03.012 (2010).

16 Nehlig, A. Effects of Coffee on the Gastro-Intestinal Tract: A Narrative Review and Literature Update. Nutrients 14, doi:10.3390/nu14020399 (2022).

17 Butt, M. S. & Sultan, M. T. Coffee and its consumption: benefits and risks. Crit Rev Food Sci Nutr 51, 363–373, doi:10.1080/10408390903586412 (2011).

18 Kang, J. et al. Increased brain volume from higher cereal and lower coffee intake: shared genetic determinants and impacts on cognition and metabolism. Cereb Cortex 32, 5163–5174, doi:10.1093/cercor/bhac005 (2022).

19 Cornelis, M. C. & van Dam, R. M. Genetic determinants of liking and intake of coffee and other bitter foods and beverages. Sci Rep 11, 23845, doi:10.1038/s41598-021-03153-7 (2021).

20 Said, M. A., van de Vegte, Y. J., Verweij, N. & van der Harst, P. Associations of Observational and Genetically Determined Caffeine Intake With Coronary Artery Disease and Diabetes Mellitus. J Am Heart Assoc 9, e016808, doi:10.1161/JAHA.120.016808 (2020).

21 Jin, T. et al. Interactions of Habitual Coffee Consumption by Genetic Polymorphisms with the Risk of Prediabetes and Type 2 Diabetes Combined. Nutrients 12, doi:10.3390/nu12082228 (2020).

22 Vogel, C. F. A., Van Winkle, L. S., Esser, C. & Haarmann-Stemmann, T. The aryl hydrocarbon receptor as a target of environmental stressors - Implications for pollution mediated stress and inflammatory responses. Redox Biol 34, 101530, doi:10.1016/j.redox.2020.101530 (2020).

23 Matoba, N. et al. GWAS of 165,084 Japanese individuals identified nine loci associated with dietary habits. Nat Hum Behav 4, 308–316, doi:10.1038/s41562-019-0805-1 (2020).

24 Kennedy, O. J. et al. Coffee Consumption and Kidney Function: A Mendelian Randomization Study. Am J Kidney Dis 75, 753–761, doi:10.1053/j.ajkd.2019.08.025 (2020).

25 Jia, H. et al. GWAS of habitual coffee consumption reveals a sex difference in the genetic effect of the 12q24 locus in the Japanese population. BMC Genet 20, 61, doi:10.1186/s12863-019-0763-7 (2019).

26 Zhong, V. W. et al. A genome-wide association study of bitter and sweet beverage consumption. Hum Mol Genet 28, 2449–2457, doi:10.1093/hmg/ddz061 (2019).

27 Nakagawa-Senda, H. et al. A genome-wide association study in the Japanese population identifies the 12q24 locus for habitual coffee consumption: The J-MICC Study. Sci Rep 8, 1493, doi:10.1038/s41598-018-19914-w (2018).

28 Cornelis, M. C. et al. Genome-wide association study of caffeine metabolites provides new insights to caffeine metabolism and dietary caffeine-consumption behavior. Hum Mol Genet 25, 5472–5482, doi:10.1093/hmg/ddw334 (2016).

29 Pirastu, N. et al. Non-additive genome-wide association scan reveals a new gene associated with habitual coffee consumption. Sci Rep 6, 31590, doi:10.1038/srep31590 (2016).

30 Coffee and Caffeine Genetics Consortium et al. Genome-wide meta-analysis identifies six novel loci associated with habitual coffee consumption. Mol Psychiatry 20, 647–656, doi:10.1038/mp.2014.107 (2015).

31 Hamza, T. H. et al. Genome-wide gene-environment study identifies glutamate receptor gene GRIN2A as a Parkinson’s disease modifier gene via interaction with coffee. PLoS Genet 7, e1002237, doi:10.1371/journal.pgen.1002237 (2011).

32 Amin, N. et al. Genome-wide association analysis of coffee drinking suggests association with CYP1A1/CYP1A2 and NRCAM. Mol Psychiatry 17, 1116–1129, doi:10.1038/mp.2011.101 (2012).

33 Sulem, P. et al. Sequence variants at CYP1A1-CYP1A2 and AHR associate with coffee consumption. Hum Mol Genet 20, 2071–2077, doi:10.1093/hmg/ddr086 (2011).

34 Yang, A., Palmer, A. A. & de Wit, H. Genetics of caffeine consumption and responses to caffeine. Psychopharmacology (Berl) 211, 245–257, doi:10.1007/s00213-010-1900-1 (2010).

35 Raja, A. M. et al. Genetic Susceptibility to Chronic Liver Disease in Individuals from Pakistan. Int J Mol Sci 21, doi:10.3390/ijms21103558 (2020).

36 Gao, H. et al. Association of GCKR Gene Polymorphisms with the Risk of Nonalcoholic Fatty Liver Disease and Coronary Artery Disease in a Chinese Northern Han Population. J Clin Transl Hepatol 7, 297–303, doi:10.14218/JCTH.2019.00030 (2019).

37 Cai, W. et al. Genetic polymorphisms associated with nonalcoholic fatty liver disease in Uyghur population: a case-control study and meta-analysis. Lipids Health Dis 18, 14, doi:10.1186/s12944-018-0877-3 (2019).

38 Xiao, X. et al. Functional POR A503V is associated with the risk of bladder cancer in a Chinese population. Sci Rep 5, 11751, doi:10.1038/srep11751 (2015).

39 Menashe, I. et al. Large-scale pathway-based analysis of bladder cancer genome-wide association data from five studies of European background. PLoS One 7, e29396, doi:10.1371/journal.pone.0029396 (2012).

40 Rotunno, M. et al. Phase I metabolic genes and risk of lung cancer: multiple polymorphisms and mRNA expression. PLoS One 4, e5652, doi:10.1371/journal.pone.0005652 (2009).

41 Aldrich, M. C. et al. CYP1A1/2 haplotypes and lung cancer and assessment of confounding by population stratification. Cancer Res 69, 2340–2348, doi:10.1158/0008-5472.CAN-08-2576 (2009).

42 Thompson, A. et al. Functional validity, role, and implications of heavy alcohol consumption genetic loci. Sci Adv 6, eaay5034, doi:10.1126/sciadv.aay5034 (2020).

43 Sanchez-Roige, S. et al. Genome-Wide Association Study Meta-Analysis of the Alcohol Use Disorders Identification Test (AUDIT) in Two Population-Based Cohorts. Am J Psychiatry 176, 107–118, doi:10.1176/appi.ajp.2018.18040369 (2019).

44 Clarke, T. K. et al. Genome-wide association study of alcohol consumption and genetic overlap with other health-related traits in UK Biobank (N=112 117). Mol Psychiatry 22, 1376–1384, doi:10.1038/mp.2017.153 (2017).

45 Yin, B., Wang, X., Huang, T. & Jia, J. Shared Genetics and Causality Between Decaffeinated Coffee Consumption and Neuropsychiatric Diseases: A Large-Scale Genome-Wide Cross-Trait Analysis and Mendelian Randomization Analysis. Front Psychiatry 13, 910432, doi:10.3389/fpsyt.2022.910432 (2022).

46 Coffee et al. Genome-wide meta-analysis identifies six novel loci associated with habitual coffee consumption. Mol Psychiatry 20, 647–656, doi:10.1038/mp.2014.107 (2015).

47 Xu, J. et al. Assessing the Association between Important Dietary Habits and Osteoporosis: A Genetic Correlation and Two-Sample Mendelian Randomization Study. Nutrients 14, doi:10.3390/nu14132656 (2022).

48 Qi, X. et al. Evaluating the Effects of Diet-Gut Microbiota Interactions on Sleep Traits Using the UK Biobank Cohort. Nutrients 14, doi:10.3390/nu14061134 (2022).

49 Wang, T. et al. Habitual coffee consumption and genetic predisposition to obesity: gene- diet interaction analyses in three US prospective studies. BMC Med 15, 97, doi:10.1186/s12916-017-0862-0 (2017).

50 Yuan, S., Daghlas, I. & Larsson, S. C. Alcohol, coffee consumption, and smoking in relation to migraine: a bidirectional Mendelian randomization study. Pain 163, e342–e348, doi:10.1097/j.pain.0000000000002360 (2022).

51 Laaboub, N. et al. Associations Between High Plasma Methylxanthine Levels, Sleep Disorders and Polygenic Risk Scores of Caffeine Consumption or Sleep Duration in a Swiss Psychiatric Cohort. Front Psychiatry 12, 756403, doi:10.3389/fpsyt.2021.756403 (2021).

52 Song, M. Y. & Park, S. Association of Polygenetic Risk Scores Related to Immunity and Inflammation with Hyperthyroidism Risk and Interactions between the Polygenetic Scores and Dietary Factors in a Large Cohort. J Thyroid Res 2021, 7664641, doi:10.1155/2021/7664641 (2021).

53 Kim, E. J. et al. Coffee Consumption and Incident Tachyarrhythmias: Reported Behavior, Mendelian Randomization, and Their Interactions. JAMA Intern Med 181, 1185–1193, doi:10.1001/jamainternmed.2021.3616 (2021).

54 Ellison-Barnes, A., Johnson, S. & Gudzune, K. Trends in Obesity Prevalence Among Adults Aged 18 Through 25 Years, 1976-2018. JAMA 326, 2073–2074, doi:10.1001/jama.2021.16685 (2021).

55 Cole, J. B., Florez, J. C. & Hirschhorn, J. N. Comprehensive genomic analysis of dietary habits in UK Biobank identifies hundreds of genetic associations. Nat Commun 11, 1467, doi:10.1038/s41467-020-15193-0 (2020).

56 Zhou, H. et al. Genome-wide meta-analysis of problematic alcohol use in 435,563 individuals yields insights into biology and relationships with other traits. Nat Neurosci 23, 809–818, doi:10.1038/s41593-020-0643-5 (2020).

57 Liu, M. et al. Association studies of up to 1.2 million individuals yield new insights into the genetic etiology of tobacco and alcohol use. Nat Genet 51, 237–244, doi:10.1038/s41588-018-0307-5 (2019).

58 Pardinas, A. F. et al. Pharmacogenomic Variants and Drug Interactions Identified Through the Genetic Analysis of Clozapine Metabolism. Am J Psychiatry 176, 477–486, doi:10.1176/appi.ajp.2019.18050589 (2019).

59 Liu, H. et al. Epigenomic and transcriptomic analyses define core cell types, genes and targetable mechanisms for kidney disease. Nat Genet 54, 950–962, doi:10.1038/s41588-022-01097-w (2022).

60 Stanzick, K. J. et al. Discovery and prioritization of variants and genes for kidney function in >1.2 million individuals. Nat Commun 12, 4350, doi:10.1038/s41467-021-24491-0 (2021).

61 Hellwege, J. N. et al. Mapping eGFR loci to the renal transcriptome and phenome in the VA Million Veteran Program. Nat Commun 10, 3842, doi:10.1038/s41467-019-11704-w (2019).

62 Wuttke, M. et al. A catalog of genetic loci associated with kidney function from analyses of a million individuals. Nat Genet 51, 957–972, doi:10.1038/s41588-019-0407-x (2019).

63 Yin, X. et al. Genome-wide association studies of metabolites in Finnish men identify disease-relevant loci. Nat Commun 13, 1644, doi:10.1038/s41467-022-29143-5 (2022).

64 Casanova, F. et al. A genome-wide association study implicates multiple mechanisms influencing raised urinary albumin-creatinine ratio. Hum Mol Genet 28, 4197–4207, doi:10.1093/hmg/ddz243 (2019).

65 Tin, A. et al. Target genes, variants, tissues and transcriptional pathways influencing human serum urate levels. Nat Genet 51, 1459–1474, doi:10.1038/s41588-019-0504-x (2019).

66 Pazoki, R. et al. GWAS for urinary sodium and potassium excretion highlights pathways shared with cardiovascular traits. Nat Commun 10, 3653, doi:10.1038/s41467-019-11451-y (2019).

67 Zanetti, D. et al. Identification of 22 novel loci associated with urinary biomarkers of albumin, sodium, and potassium excretion. Kidney Int 95, 1197–1208, doi:10.1016/j.kint.2018.12.017 (2019).

68 Haas, M. E. et al. Genetic Association of Albuminuria with Cardiometabolic Disease and Blood Pressure. Am J Hum Genet 103, 461–473, doi:10.1016/j.ajhg.2018.08.004 (2018).

69 Jimenez, A., Adisa, A., Woodham, C. & Saleh, M. Determination of polycyclic aromatic hydrocarbons in roasted coffee. J Environ Sci Health B 49, 828–835, doi:10.1080/03601234.2014.938552 (2014).

70 Teumer, A. et al. Genome-wide association meta-analyses and fine-mapping elucidate pathways influencing albuminuria. Nat Commun 10, 4130, doi:10.1038/s41467-019-11576-0 (2019).

71 Fredholm, B. B., Chen, J. F., Masino, S. A. & Vaugeois, J. M. Actions of adenosine at its receptors in the CNS: insights from knockouts and drugs. Annu Rev Pharmacol Toxicol 45, 385–412, doi:10.1146/annurev.pharmtox.45.120403.095731 (2005).

72 Tanner, J. A. & Tyndale, R. F. Variation in CYP2A6 Activity and Personalized Medicine. J Pers Med 7, doi:10.3390/jpm7040018 (2017).

73 Saunders, G. R. B. et al. Genetic diversity fuels gene discovery for tobacco and alcohol use. Nature 612, 720–724, doi:10.1038/s41586-022-05477-4 (2022).

74 Sakaue, S. et al. A cross-population atlas of genetic associations for 220 human phenotypes. Nat Genet 53, 1415–1424, doi:10.1038/s41588-021-00931-x (2021).

75 Koskeridis, F. et al. Pleiotropic genetic architecture and novel loci for C-reactive protein levels. Nat Commun 13, 6939, doi:10.1038/s41467-022-34688-6 (2022).

76 Deak, J. D. et al. Genome-wide association study in individuals of European and African ancestry and multi-trait analysis of opioid use disorder identifies 19 independent genome-wide significant risk loci. Mol Psychiatry 27, 3970–3979, doi:10.1038/s41380-022-01709-1 (2022).

77 Pasman, J. A. et al. Genetic Risk for Smoking: Disentangling Interplay Between Genes and Socioeconomic Status. Behav Genet 52, 92–107, doi:10.1007/s10519-021-10094-4 (2022).

78 Xu, K. et al. Genome-wide association study of smoking trajectory and meta-analysis of smoking status in 842,000 individuals. Nat Commun 11, 5302, doi:10.1038/s41467-020-18489-3 (2020).

79 Pasman, J. A. et al. GWAS of lifetime cannabis use reveals new risk loci, genetic overlap with psychiatric traits, and a causal influence of schizophrenia. Nat Neurosci 21, 1161–1170, doi:10.1038/s41593-018-0206-1 (2018).

80 Schumann, G. et al. KLB is associated with alcohol drinking, and its gene product beta- Klotho is necessary for FGF21 regulation of alcohol preference. Proc Natl Acad Sci U S A 113, 14372–14377, doi:10.1073/pnas.1611243113 (2016).

81 Stringer, S. et al. Genome-wide association study of lifetime cannabis use based on a large meta-analytic sample of 32 330 subjects from the International Cannabis Consortium. Transl Psychiatry 6, e769, doi:10.1038/tp.2016.36 (2016).

82 Wain, L. V. et al. Novel insights into the genetics of smoking behaviour, lung function, and chronic obstructive pulmonary disease (UK BiLEVE): a genetic association study in UK Biobank. Lancet Respir Med 3, 769–781, doi:10.1016/S2213-2600(15)00283-0 (2015).

83 King, C. P. et al. Cdh13 and AdipoQ gene knockout alter instrumental and Pavlovian drug conditioning. Genes Brain Behav 16, 686–698, doi:10.1111/gbb.12382 (2017).

84 Drgonova, J. et al. Cadherin 13: human cis-regulation and selectively-altered addiction phenotypes and cerebral cortical dopamine in knockout mice. Mol Med 22, 537–547, doi:10.2119/molmed.2015.00170 (2016).

85 Yang, J. et al. The contribution of rare and common variants in 30 genes to risk nicotine dependence. Mol Psychiatry 20, 1467–1478, doi:10.1038/mp.2014.156 (2015).

86 Weber, M. et al. Increased polysialic acid neural cell adhesion molecule expression in human hippocampus of heroin addicts. Neuroscience 138, 1215–1223, doi:10.1016/j.neuroscience.2005.11.059 (2006).

87 Barker, J. M., Torregrossa, M. M. & Taylor, J. R. Low prefrontal PSA-NCAM confers risk for alcoholism-related behavior. Nat Neurosci 15, 1356–1358, doi:10.1038/nn.3194 (2012).

88 Mackowiak, M. et al. Cocaine decreases the expression of PSA-NCAM protein and attenuates long-term potentiation via glucocorticoid receptors in the rat dentate gyrus. Eur J Neurosci 27, 2928–2937, doi:10.1111/j.1460-9568.2008.06255.x (2008).

89 Kojetin, D. J. & Burris, T. P. REV-ERB and ROR nuclear receptors as drug targets. Nat Rev Drug Discov 13, 197–216, doi:10.1038/nrd4100 (2014).

90 Lee, J. J. et al. Gene discovery and polygenic prediction from a genome-wide association study of educational attainment in 1.1 million individuals. Nat Genet 50, 1112–1121, doi:10.1038/s41588-018-0147-3 (2018).

91 Chen, M. H. et al. Trans-ethnic and Ancestry-Specific Blood-Cell Genetics in 746,667 Individuals from 5 Global Populations. Cell 182, 1198–1213 e1114, doi:10.1016/j.cell.2020.06.045 (2020).

92 Thompson, A., King, K., Morris, A. P. & Pirmohamed, M. Assessing the impact of alcohol consumption on the genetic contribution to mean corpuscular volume. Hum Mol Genet 30, 2040–2051, doi:10.1093/hmg/ddab147 (2021).

93 Willer, C. J., Li, Y. & Abecasis, G. R. METAL: fast and efficient meta-analysis of genomewide association scans. Bioinformatics 26, 2190–2191, doi:10.1093/bioinformatics/btq340 (2010).

94 Zhu, Z. et al. Causal associations between risk factors and common diseases inferred from GWAS summary data. Nat Commun 9, 224, doi:10.1038/s41467-017-02317-2 (2018).

95 Kendler, K. S., Schmitt, E., Aggen, S. H. & Prescott, C. A. Genetic and environmental influences on alcohol, caffeine, cannabis, and nicotine use from early adolescence to middle adulthood. Arch Gen Psychiatry 65, 674–682, doi:10.1001/archpsyc.65.6.674 (2008).

96 Karlsson Linner, R., et al. Multivariate analysis of 1.5 million people identifies genetic associations with traits related to self-regulation and addiction. Nat Neurosci 24, 1367–1376, doi:10.1038/s41593-021-00908-3 (2021).

97 Wojcik, G. L. et al. Genetic analyses of diverse populations improves discovery for complex traits. Nature 570, 514–518, doi:10.1038/s41586-019-1310-4 (2019).

98 Vuckovic, D. et al. The Polygenic and Monogenic Basis of Blood Traits and Diseases. Cell 182, 1214–1231 e1211, doi:10.1016/j.cell.2020.08.008 (2020).

99 Klarin, D. et al. Genetics of blood lipids among ∼300,000 multi-ethnic participants of the Million Veteran Program. Nat Genet 50, 1514–1523, doi:10.1038/s41588-018-0222-9 (2018).

100 Feitosa, M. F. et al. Novel genetic associations for blood pressure identified via gene- alcohol interaction in up to 570K individuals across multiple ancestries. PLoS One 13, e0198166, doi:10.1371/journal.pone.0198166 (2018).

101 Sung, Y. J. et al. A Large-Scale Multi-ancestry Genome-wide Study Accounting for Smoking Behavior Identifies Multiple Significant Loci for Blood Pressure. Am J Hum Genet 102, 375–400, doi:10.1016/j.ajhg.2018.01.015 (2018).

102 Astle, W. J. et al. The Allelic Landscape of Human Blood Cell Trait Variation and Links to Common Complex Disease. Cell 167, 1415–1429 e1419, doi:10.1016/j.cell.2016.10.042 (2016).

103 Meeks, K. A. C. et al. Genome-wide analyses of multiple obesity-related cytokines and hormones informs biology of cardiometabolic traits. Genome Med 13, 156, doi:10.1186/s13073-021-00971-2 (2021).

104 German, C. A., Sinsheimer, J. S., Klimentidis, Y. C., Zhou, H. & Zhou, J. J. Ordered multinomial regression for genetic association analysis of ordinal phenotypes at Biobank scale. Genet Epidemiol 44, 248–260, doi:10.1002/gepi.22276 (2020).

105 Pham, K. et al. High coffee consumption, brain volume and risk of dementia and stroke. Nutr Neurosci 25, 2111–2122, doi:10.1080/1028415X.2021.1945858 (2022).

106 Haller, S., Montandon, M. L., Rodriguez, C., Herrmann, F. R. & Giannakopoulos, P. Impact of Coffee, Wine, and Chocolate Consumption on Cognitive Outcome and MRI Parameters in Old Age. Nutrients 10, doi:10.3390/nu10101391 (2018).

107 Araujo, L. F. et al. Association of Coffee Consumption with MRI Markers and Cognitive Function: A Population-Based Study. J Alzheimers Dis 53, 451–461, doi:10.3233/JAD-160116 (2016).

108 Perlaki, G. et al. Coffee consumption may influence hippocampal volume in young women. Brain Imaging Behav 5, 274–284, doi:10.1007/s11682-011-9131-6 (2011).

109 Magalhaes, R. et al. Habitual coffee drinkers display a distinct pattern of brain functional connectivity. Mol Psychiatry 26, 6589–6598, doi:10.1038/s41380-021-01075-4 (2021).

110 Laurienti, P. J. et al. Dietary caffeine consumption modulates fMRI measures. Neuroimage 17, 751–757 (2002).

111 Deak, J. D. & Johnson, E. C. Genetics of substance use disorders: a review. Psychol Med 51, 2189–2200, doi:10.1017/S0033291721000969 (2021).

112 Vanyukov, M. M. et al. Common liability to addiction and “gateway hypothesis”: theoretical, empirical and evolutionary perspective. Drug Alcohol Depend 123 **Suppl 1**, S3–17, doi:10.1016/j.drugalcdep.2011.12.018 (2012).

113 Chang, L. H. et al. Investigating the genetic and causal relationship between initiation or use of alcohol, caffeine, cannabis and nicotine. Drug Alcohol Depend 210, 107966, doi:10.1016/j.drugalcdep.2020.107966 (2020).

114 Hettema, J. M., Corey, L. A. & Kendler, K. S. A multivariate genetic analysis of the use of tobacco, alcohol, and caffeine in a population based sample of male and female twins. Drug Alcohol Depend 57, 69–78, doi:10.1016/s0376-8716(99)00053-8 (1999).

115 Swan, G. E., Carmelli, D. & Cardon, L. R. The consumption of tobacco, alcohol, and coffee in Caucasian male twins: a multivariate genetic analysis. J Subst Abuse 8, 19–31, doi:10.1016/s0899-3289(96)90055-3 (1996).

116 Mallard, T. T. & Sanchez-Roige, S. Dimensional Phenotypes in Psychiatric Genetics: Lessons from Genome-Wide Association Studies of Alcohol Use Phenotypes. Complex Psychiatry 7, 45–48, doi:10.1159/000518863 (2021).

117 Gelernter, J. & Polimanti, R. Genetics of substance use disorders in the era of big data. Nat Rev Genet 22, 712–729, doi:10.1038/s41576-021-00377-1 (2021).

118 Johnson, E. C. et al. A large-scale genome-wide association study meta-analysis of cannabis use disorder. Lancet Psychiatry 7, 1032–1045, doi:10.1016/S2215-0366(20)30339-4 (2020).

119 Sanchez-Roige, S., Palmer, A. A. & Clarke, T. K. Recent Efforts to Dissect the Genetic Basis of Alcohol Use and Abuse. Biol Psychiatry 87, 609–618, doi:10.1016/j.biopsych.2019.09.011 (2020).

120 Ramli, N. N. S., Alkhaldy, A. A. & Mhd Jalil, A. M. Effects of Caffeinated and Decaffeinated Coffee Consumption on Metabolic Syndrome Parameters: A Systematic Review and Meta-Analysis of Data from Randomised Controlled Trials. Medicina (Kaunas) 57, doi:10.3390/medicina57090957 (2021).

121 Lee, A. et al. Coffee Intake and Obesity: A Meta-Analysis. Nutrients 11, doi:10.3390/nu11061274 (2019).

122 Ludwig, I. A. et al. Variations in caffeine and chlorogenic acid contents of coffees: what are we drinking? Food Funct 5, 1718–1726, doi:10.1039/c4fo00290c (2014).

123 Schwartz, A. & Bellissimo, N. Nicotine and energy balance: A review examining the effect of nicotine on hormonal appetite regulation and energy expenditure. Appetite 164, 105260, doi:10.1016/j.appet.2021.105260 (2021).

124 Schubert, M. M. et al. Caffeine, coffee, and appetite control: a review. Int J Food Sci Nutr 68, 901–912, doi:10.1080/09637486.2017.1320537 (2017).

125 Cornelis, M. C., Bennett, D. A., Weintraub, S., Schneider, J. A. & Morris, M. C. Caffeine Consumption and Dementia: Are Lewy Bodies the Link? Ann Neurol 91, 834–846, doi:10.1002/ana.26349 (2022).

126 Di Maso, M., Boffetta, P., Negri, E., La Vecchia, C. & Bravi, F. Caffeinated Coffee Consumption and Health Outcomes in the US Population: A Dose-Response Meta- Analysis and Estimation of Disease Cases and Deaths Avoided. Adv Nutr 12, 1160–1176, doi:10.1093/advances/nmaa177 (2021).

127 Mentis, A. A., Dardiotis, E., Efthymiou, V. & Chrousos, G. P. Non-genetic risk and protective factors and biomarkers for neurological disorders: a meta-umbrella systematic review of umbrella reviews. BMC Med 19, 6, doi:10.1186/s12916-020-01873-7 (2021).

128 Chen, X., Zhao, Y., Tao, Z. & Wang, K. Coffee consumption and risk of prostate cancer: a systematic review and meta-analysis. BMJ Open 11, e038902, doi:10.1136/bmjopen-2020-038902 (2021).

129 Hong, C. T., Chan, L. & Bai, C. H. The Effect of Caffeine on the Risk and Progression of Parkinson’s Disease: A Meta-Analysis. Nutrients 12, doi:10.3390/nu12061860 (2020).

130 Sartini, M. et al. Coffee Consumption and Risk of Colorectal Cancer: A Systematic Review and Meta-Analysis of Prospective Studies. Nutrients 11, doi:10.3390/nu11030694 (2019).

131 Loftfield, E. et al. Association of Coffee Drinking With Mortality by Genetic Variation in Caffeine Metabolism: Findings From the UK Biobank. JAMA Intern Med 178, 1086–1097, doi:10.1001/jamainternmed.2018.2425 (2018).

132 Wu, L., Sun, D. & He, Y. Coffee intake and the incident risk of cognitive disorders: A dose-response meta-analysis of nine prospective cohort studies. Clin Nutr 36, 730–736, doi:10.1016/j.clnu.2016.05.015 (2017).

133 Liu, Q. P. et al. Habitual coffee consumption and risk of cognitive decline/dementia: A systematic review and meta-analysis of prospective cohort studies. Nutrition 32, 628–636, doi:10.1016/j.nut.2015.11.015 (2016).

134 Kennedy, O. J. et al. Systematic review with meta-analysis: coffee consumption and the risk of cirrhosis. Aliment Pharmacol Ther 43, 562–574, doi:10.1111/apt.13523 (2016).

135 Crippa, A., Discacciati, A., Larsson, S. C., Wolk, A. & Orsini, N. Coffee consumption and mortality from all causes, cardiovascular disease, and cancer: a dose-response meta- analysis. Am J Epidemiol 180, 763–775, doi:10.1093/aje/kwu194 (2014).

136 Ding, M., Bhupathiraju, S. N., Satija, A., van Dam, R. M. & Hu, F. B. Long-term coffee consumption and risk of cardiovascular disease: a systematic review and a dose- response meta-analysis of prospective cohort studies. Circulation 129, 643–659, doi:10.1161/CIRCULATIONAHA.113.005925 (2014).

137 Abalo, R. Coffee and Caffeine Consumption for Human Health. Nutrients 13, doi:10.3390/nu13092918 (2021).

138 Hou, C. et al. Medical conditions associated with coffee consumption: Disease-trajectory and comorbidity network analyses of a prospective cohort study in UK Biobank. Am J Clin Nutr 116, 730–740, doi:10.1093/ajcn/nqac148 (2022).

139 Paz-Graniel, I. et al. Association between coffee consumption and total dietary caffeine intake with cognitive functioning: cross-sectional assessment in an elderly Mediterranean population. Eur J Nutr 60, 2381–2396, doi:10.1007/s00394-020-02415-w (2021).

140 Dong, X., Li, S., Sun, J., Li, Y. & Zhang, D. Association of Coffee, Decaffeinated Coffee and Caffeine Intake from Coffee with Cognitive Performance in Older Adults: National Health and Nutrition Examination Survey (NHANES) 2011-2014. Nutrients 12, doi:10.3390/nu12030840 (2020).

141 Haskell-Ramsay, C. F. et al. The Acute Effects of Caffeinated Black Coffee on Cognition and Mood in Healthy Young and Older Adults. Nutrients 10, doi:10.3390/nu10101386 (2018).

142 Johnson-Kozlow, M., Kritz-Silverstein, D., Barrett-Connor, E. & Morton, D. Coffee consumption and cognitive function among older adults. Am J Epidemiol 156, 842–850, doi:10.1093/aje/kwf119 (2002).

143 Kim, Y., Je, Y. & Giovannucci, E. Coffee consumption and all-cause and cause-specific mortality: a meta-analysis by potential modifiers. Eur J Epidemiol 34, 731–752, doi:10.1007/s10654-019-00524-3 (2019).

144 Abdellaoui, A., Dolan, C. V., Verweij, K. J. H. & Nivard, M. G. Gene-environment correlations across geographic regions affect genome-wide association studies. Nat Genet 54, 1345–1354, doi:10.1038/s41588-022-01158-0 (2022).

145 De Toni, L. et al. Phthalates and heavy metals as endocrine disruptors in food: A study on pre-packed coffee products. Toxicol Rep 4, 234–239, doi:10.1016/j.toxrep.2017.05.004 (2017).

146 Sakaki, J. R., Melough, M. M., Provatas, A. A., Perkins, C. & Chun, O. K. Evaluation of estrogenic chemicals in capsule and French press coffee using ultra-performance liquid chromatography with tandem mass spectrometry. Toxicol Rep 7, 1020–1024, doi:10.1016/j.toxrep.2020.08.015 (2020).

147 Chieng, D. et al. The impact of coffee subtypes on incident cardiovascular disease, arrhythmias, and mortality: long-term outcomes from the UK Biobank. Eur J Prev Cardiol 29, 2240–2249, doi:10.1093/eurjpc/zwac189 (2022).

148 Nordestgaard, A. T. Causal relationship from coffee consumption to diseases and mortality: a review of observational and Mendelian randomization studies including cardiometabolic diseases, cancer, gallstones and other diseases. Eur J Nutr 61, 573–587, doi:10.1007/s00394-021-02650-9 (2022).

149 Treur, J. L. et al. Smoking and caffeine consumption: a genetic analysis of their association. Addict Biol 22, 1090–1102, doi:10.1111/adb.12391 (2017).

150 Verweij, K. J. H., Treur, J. L. & Vink, J. M. Investigating causal associations between use of nicotine, alcohol, caffeine and cannabis: a two-sample bidirectional Mendelian randomization study. Addiction 113, 1333–1338, doi:10.1111/add.14154 (2018).

151 Ware, J. J. et al. Does coffee consumption impact on heaviness of smoking? Addiction 112, 1842–1853, doi:10.1111/add.13888 (2017).

152 Sanchez-Roige, S. & Palmer, A. A. Emerging phenotyping strategies will advance our understanding of psychiatric genetics. Nat Neurosci 23, 475–480, doi:10.1038/s41593-020-0609-7 (2020).

153 Jee, H. J., Lee, S. G., Bormate, K. J. & Jung, Y. S. Effect of Caffeine Consumption on the Risk for Neurological and Psychiatric Disorders: Sex Differences in Human. Nutrients 12, doi:10.3390/nu12103080 (2020).

154 Nehlig, A. Interindividual Differences in Caffeine Metabolism and Factors Driving Caffeine Consumption. Pharmacol Rev 70, 384–411, doi:10.1124/pr.117.014407 (2018).

155 Sanchez-Roige, S. et al. CADM2 is implicated in impulsive personality traits by genome- and phenome-wide association studies in humans, with further support from studies of Cadm2 mutant mice. Translational Psychiary 13, 167, doi:10.1038/s41398-023-02453-y (2023).

156 Durand, E. Y., Do, C. B., Mountain, J. L. & Macpherson, J. M. Ancestry Composition: A Novel, Efficient Pipeline for Ancestry Deconvolution. bioRxiv. doi:10.1101/010512 (2014).

157 Hyde, C. L. et al. Identification of 15 genetic loci associated with risk of major depression in individuals of European descent. Nat Genet 48, 1031–1036, doi:10.1038/ng.3623 (2016).

158 Eriksson, N. et al. Web-based, participant-driven studies yield novel genetic associations for common traits. PLoS Genet 6, e1000993, doi:10.1371/journal.pgen.1000993 (2010).

159 Lam, M. et al. Large-Scale Cognitive GWAS Meta-Analysis Reveals Tissue-Specific Neural Expression and Potential Nootropic Drug Targets. Cell Rep 21, 2597–2613, doi:10.1016/j.celrep.2017.11.028 (2017).

160 Sanchez-Roige, S. et al. Genome-wide association study of delay discounting in 23,217 adult research participants of European ancestry. Nat Neurosci 21, 16–18, doi:10.1038/s41593-017-0032-x (2018).

161 Sanchez-Roige, S. et al. Genome-wide association study of problematic opioid prescription use in 132,113 23andMe research participants of European ancestry. Mol Psychiatry 26, 6209–6217, doi:10.1038/s41380-021-01335-3 (2021).

162 Sey, N. Y. A. et al. A computational tool (H-MAGMA) for improved prediction of brain- disorder risk genes by incorporating brain chromatin interaction profiles. Nat Neurosci 23, 583–593, doi:10.1038/s41593-020-0603-0 (2020).

163 Barbeira, A. N. et al. Exploring the phenotypic consequences of tissue specific gene expression variation inferred from GWAS summary statistics. Nat Commun 9, 1825, doi:10.1038/s41467-018-03621-1 (2018).

164 Barbeira, A. N. et al. Integrating predicted transcriptome from multiple tissues improves association detection. PLoS Genet 15, e1007889, doi:10.1371/journal.pgen.1007889 (2019).

165 Bulik-Sullivan, B. K. et al. LD Score regression distinguishes confounding from polygenicity in genome-wide association studies. Nat Genet 47, 291–295, doi:10.1038/ng.3211 (2015).

166 Meddens, S. F. W. et al. Genomic analysis of diet composition finds novel loci and associations with health and lifestyle. Mol Psychiatry 26, 2056–2069, doi:10.1038/s41380-020-0697-5 (2021).

167 Ge, T., Chen, C. Y., Ni, Y., Feng, Y. A. & Smoller, J. W. Polygenic prediction via Bayesian regression and continuous shrinkage priors. Nat Commun 10, 1776, doi:10.1038/s41467-019-09718-5 (2019).

168 Roden, D. M. et al. Development of a large-scale de-identified DNA biobank to enable personalized medicine. Clin Pharmacol Ther 84, 362–369, doi:10.1038/clpt.2008.89 (2008).

169 Dennis, J. et al. Genetic risk for major depressive disorder and loneliness in sex-specific associations with coronary artery disease. Mol Psychiatry 26, 4254–4264, doi:10.1038/s41380-019-0614-y (2021).

170 Carroll, R. J., Bastarache, L. & Denny, J. C. R PheWAS: data analysis and plotting tools for phenome-wide association studies in the R environment. Bioinformatics 30, 2375–2376, doi:10.1093/bioinformatics/btu197 (2014).

171 Kaufman, L. & Rousseeuw, P. J. Finding groups in data : an introduction to cluster analysis. (Wiley, 1990).

172 Watanabe, K. et al. A global overview of pleiotropy and genetic architecture in complex traits. Nat Genet 51, 1339–1348, doi:10.1038/s41588-019-0481-0 (2019).

173 Walters, R. K. et al. Transancestral GWAS of alcohol dependence reveals common genetic underpinnings with psychiatric disorders. Nat Neurosci 21, 1656–1669, doi:10.1038/s41593-018-0275-1 (2018).

174 Quach, B. C. et al. Expanding the genetic architecture of nicotine dependence and its shared genetics with multiple traits. Nat Commun 11, 5562, doi:10.1038/s41467-020-19265-z (2020).

175 Karlsson Linner, R., et al. Genome-wide association analyses of risk tolerance and risky behaviors in over 1 million individuals identify hundreds of loci and shared genetic influences. Nat Genet 51, 245–257, doi:10.1038/s41588-018-0309-3 (2019).

176 Barkley-Levenson, A. M., Lagarda, F. A. & Palmer, A. A. Glyoxalase 1 (GLO1) Inhibition or Genetic Overexpression Does Not Alter Ethanol’s Locomotor Effects: Implications for GLO1 as a Therapeutic Target in Alcohol Use Disorders. Alcohol Clin Exp Res 42, 869–878, doi:10.1111/acer.13623 (2018).

177 Distler, M. G. et al. Glyoxalase 1 increases anxiety by reducing GABAA receptor agonist methylglyoxal. J Clin Invest 122, 2306–2315, doi:10.1172/JCI61319 (2012).

178 Dawson, G. R. & Tricklebank, M. D. Use of the elevated plus maze in the search for novel anxiolytic agents. Trends Pharmacol Sci 16, 33–36, doi:10.1016/s0165-6147(00)88973-7 (1995).

179 Henn, B. M. et al. Cryptic distant relatives are common in both isolated and cosmopolitan genetic samples. PLoS One 7, e34267, doi:10.1371/journal.pone.0034267 (2012).

180 Fuchsberger, C., Abecasis, G. R. & Hinds, D. A. minimac2: faster genotype imputation. Bioinformatics 31, 782–784, doi:10.1093/bioinformatics/btu704 (2015).

181 Pruim, R. J. et al. LocusZoom: regional visualization of genome-wide association scan results. Bioinformatics 26, 2336–2337, doi:10.1093/bioinformatics/btq419 (2010).

