## Supplementary Material for "Genome-Wide Association Studies of Coffee Intake in UK/US Participants of European Ancestry Uncover Gene-Cohort Influences"

**Analysis of local ancestry**

Ancestry was determined by analyzing local ancestry^1^. Briefly, the 23andMe algorithm first partitions phased genomic data into short windows of about 300 SNPs. Within each window, we use a support vector machine to classify individual haplotypes into one of 31 reference populations (<https://www.23andme.com/ancestry-composition-guide/>). The support vector machine classifications are fed into a hidden Markov model that accounts for switch errors and incorrect assignments, and gives probabilities for each reference population in each window. Finally, we used simulated admixed individuals to recalibrate the hidden Markov model probabilities so that the reported assignments are consistent with the simulated admixture proportions. The reference population data is derived from public datasets (the Human Genome Diversity Project, HapMap, and 1000 Genomes), as well as 23andMe customers who have reported having four grandparents from the same country.

Ancestries are defined as follows:

| **Ancestry** | **Classification Criteria** |
| --- | --- |
| European | European + Middle Eastern > 0.97, European > 0.90 |
| East Asian | East Asian + Southeast Asian > 0.97 |
| South Asian | South Asian > 0.97 |
| Middle Eastern (& North African) | Middle Eastern + European > 0.97, Middle Eastern > 0.90 |
| African American + Latin American | European + African + East Asian + Native American + Middle Eastern > 0.90, African + Native American > 0.01 |

African Americans and Latin American are admixed with broadly varying contributions from Europe, Africa and the Americas. The distributions of the length of segments of European, African and American ancestry are very different between African Americans and Latin American because of distinct admixture timing between the three ancestral populations in the two ethnic groups. Therefore, we trained a logistic classifier that takes one individual's length histogram of segments of African, European and American ancestry and predicts whether the customer is likely African American or Latin American.

**Genome-wide association and secondary analyses**

Participants were genotyped on one of five Illumina genotyping platforms, containing between 550,000 to 950,000 variants, for a total of 1.6 million genotyped variants. Samples that failed to reach 98.5% call rate were re-analyzed. Genotyping quality controls included discarding variants with a Hardy-Weinberg *p<*1.00E−20, batch effects (ANOVA *p<*1.00E-20), or a call rate of <90%^2-4^. About 64.4M variants were then imputed against the Haplotype Reference Consortium (**HRC**) panel, augmented by a single unified imputation reference panel combining the May 2015 release of the 1000 Genomes Phase 3 haplotypes with the UK10K imputation reference panel for variants not present in the HRC. Imputed variants with low imputation quality (r^2^<0.50 averaged across batches or a minimum r^2^<0.30), or with evidence of batch effects (p<1.00E-50) were removed^2,3^. A total of 1.3M genotyped and 30.5M imputed variants passed the pre- and post-GWAS quality controls. We furthermore filtered out variants with minor allele frequency (**MAF**) <0.1%, which are extremely sensitive to quantitative trait over-dispersion, reducing to 14.1M variants available for follow-up analyses. Principal components were computed using ~65,000 high-quality genotyped variants present in all five genotyping platforms.

A maximal set of unrelated individuals was chosen for the analysis using a segmental identity-by-descent (**IBD**) estimation algorithm^5^ to ensure that only unrelated individuals were included in the sample. Individuals were defined as related if they shared more than 700 cM IBD, including regions where the two individuals shared either one or both genomic segments IBD. This level of relatedness (~20% of the genome) corresponds to approximately the minimal expected sharing between first cousins in an outbred population.

We imputed participant genotype data against the September 2013 release of 1000 Genomes phase 1 version 3 reference haplotypes. We phased and imputed data for each genotyping platform separately. We phased using an internally developed phasing tool, Finch, which implements the Beagle haplotype graph-based phasing algorithm, modified to separate the haplotype graph construction and phasing steps. In preparation for imputation, we split phased chromosomes into segments of no more than 10,000 genotyped SNPs, with overlaps of 200 SNPs. We excluded SNPs with Hardy-Weinberg equilibrium *p<*10E-20, call rate <95%, or with large allele frequency discrepancies compared to European 1000 Genomes reference data. Frequency discrepancies were identified by computing a 2 x 2 table of allele counts for European 1000 Genomes samples and 2000 randomly sampled 23andMe research participants with European ancestry, and identifying SNPs with a 𝜒^2^ *p<*10E-15. We imputed each phased segment against all-ethnicity 1000 Genomes haplotypes (excluding monomorphic and singleton sites) using Minimac2^6^, using 5 rounds and 200 states for parameter estimation.

For the X chromosome, we built separate haplotype graphs for the non-pseudoautosomal region and each pseudoautosomal region, and these regions were phased separately. We then imputed males and females together using Minimac2, as with the autosomes, treating males as homozygous pseudo-diploids for the non-pseudoautosomal region.

For tests using imputed data, we use the imputed dosages rather than best-guess genotypes. We imputed HLA allele dosages from SNP genotype data using HIBAG. We imputed alleles for HLA-A, B, C, DPB1, DQA1, DQB1, and DRB1 loci at four-digit resolution. To test associations between HLA allele dosages and phenotypes, we performed linear regression using the same set of covariates used in the SNP based GWAS. We performed separate association tests for each imputed allele. HLA analysis did not identify any significant signal and hence are not described in the main text.

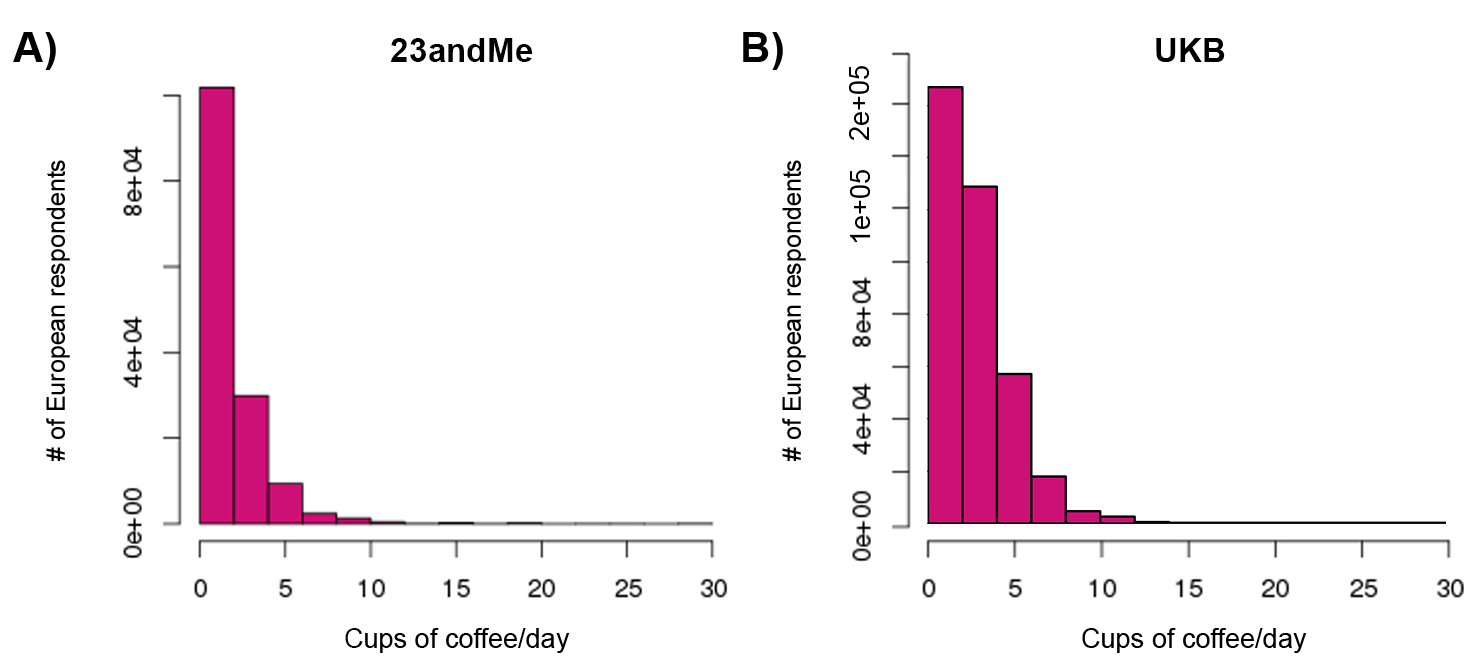

**Supplementary Figure 1.** Self-reported daily **A)** caffeinated coffee intake among 23andMe and **B)** any coffee intake among UKB participants.

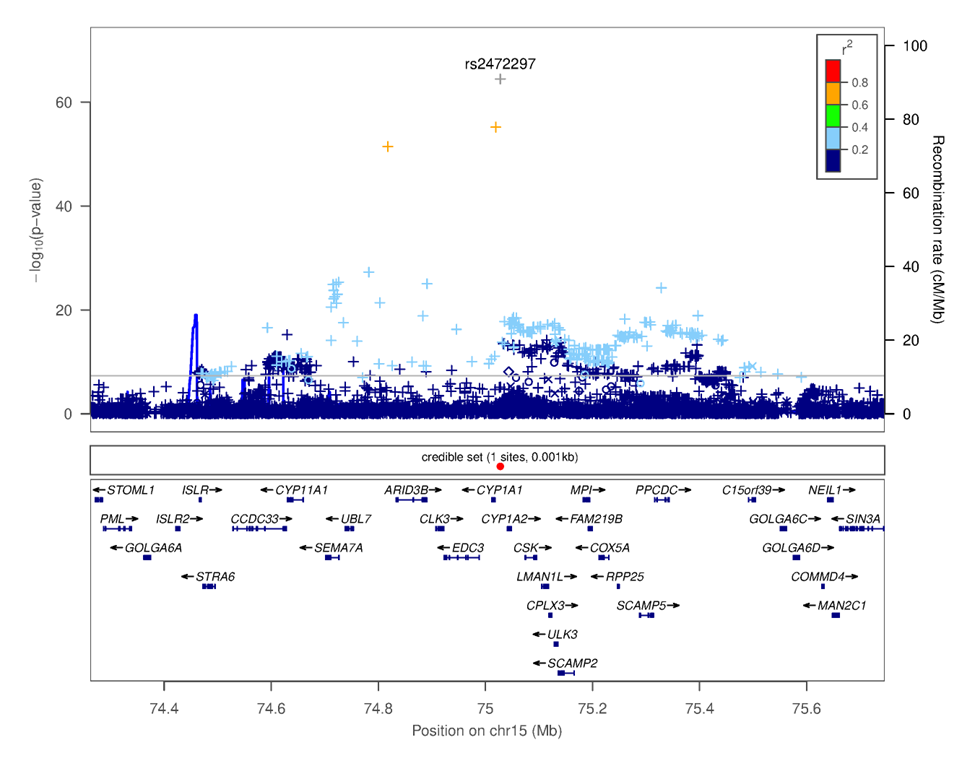

**Supplementary Figure 2.** Locus zoom plot focusing on SNP rs2472297 on chromosome 15. This plot was generated using LocusZoom^7^. The -log_10_(*p-*value) is shown on the left *y*-axis; position in Mb is on the *x*-axis. Recombination rates (expressed in centiMorgans cM per Mb; NCBI Build GRCh37; highlighted in blue) are shown on the right *y*-axis. Pairwise linkage disequilibrium (r^2^) of each SNP with the top SNP in the region is indicated by its color. Crossed points represent imputed SNPs, circles represent directly genotyped SNPs.

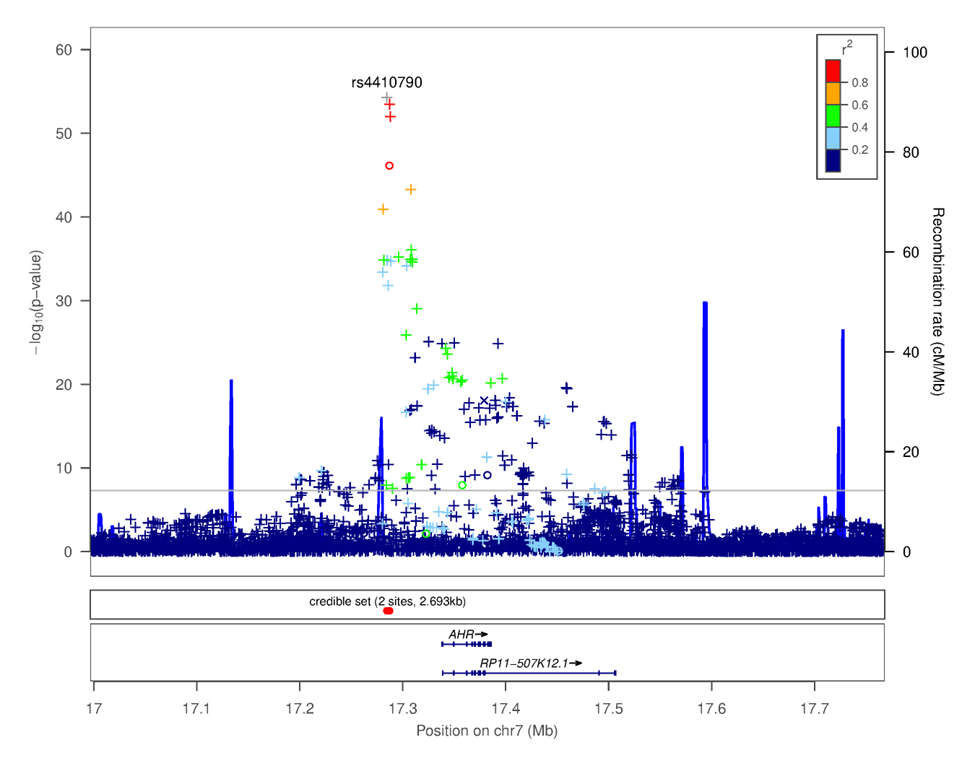
**Supplementary Figure 3.** Locus zoom plot focusing on SNP rs4410790 on chromosome 7. This plot was generated using LocusZoom^7^. The -log_10_(*p-*value) is shown on the left *y*-axis; position in Mb is on the *x*-axis. Recombination rates (expressed in centiMorgans cM per Mb; NCBI Build GRCh37; highlighted in blue) are shown on the right *y*-axis. Pairwise linkage disequilibrium (r^2^) of each SNP with the top SNP in the region is indicated by its color. Crossed points represent imputed SNPs, circles represent directly genotyped SNPs.

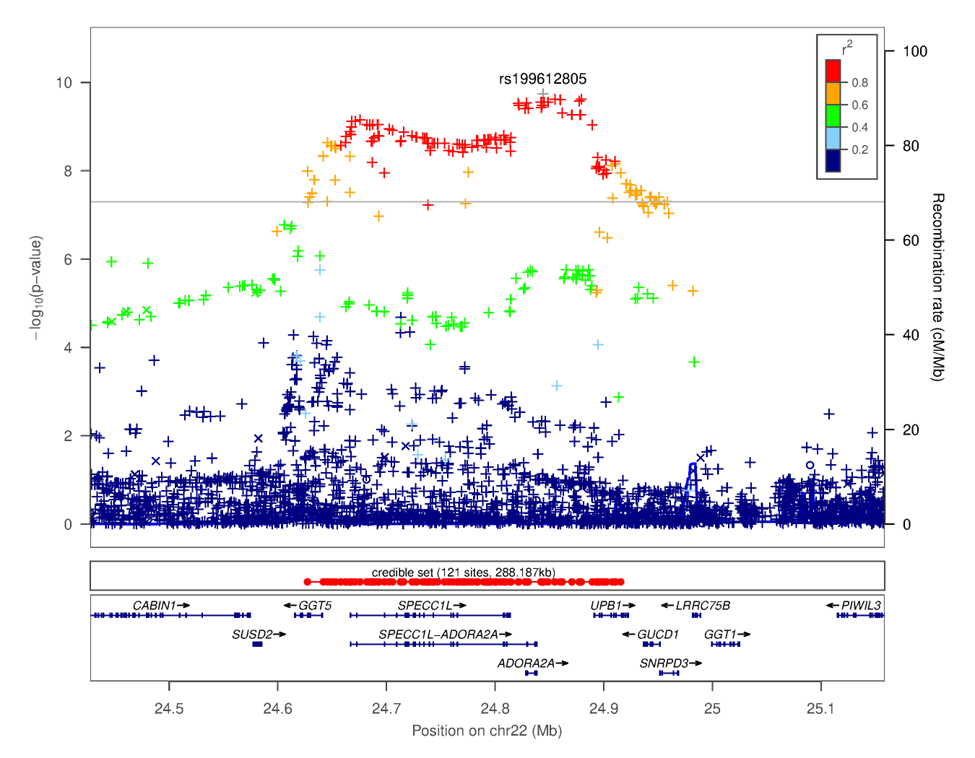

**Supplementary Figure 4.** Locus zoom plot focusing on SNP rs199612805 on chromosome 22. This plot was generated using LocusZoom^7^. The -log_10_(*p-*value) is shown on the left *y*-axis; position in Mb is on the *x*-axis. Recombination rates (expressed in centiMorgans cM per Mb; NCBI Build GRCh37; highlighted in blue) are shown on the right *y*-axis. Pairwise linkage disequilibrium (r^2^) of each SNP with the top SNP in the region is indicated by its color. Crossed points represent imputed SNPs, circles represent directly genotyped SNPs.

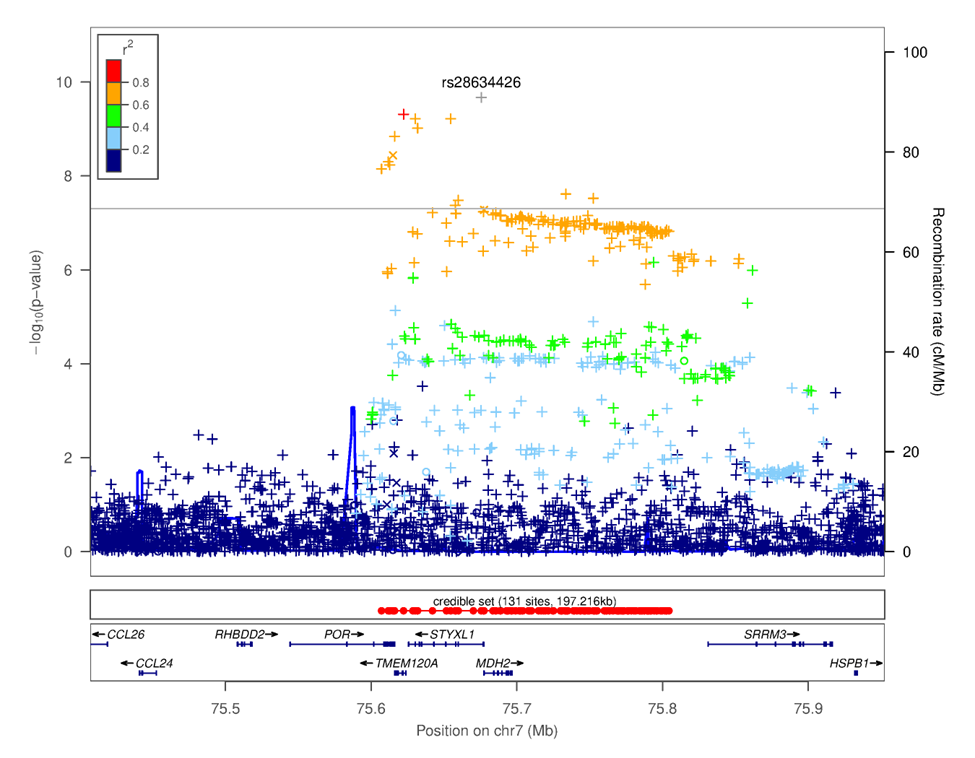

**Supplementary Figure 5.** Locus zoom plot focusing on SNP rs28634426 on chromosome 7. This plot was generated using LocusZoom^7^. The -log_10_(*p-*value) is shown on the left *y*-axis; position in Mb is on the *x*-axis. Recombination rates (expressed in centiMorgans cM per Mb; NCBI Build GRCh37; highlighted in blue) are shown on the right *y*-axis. Pairwise linkage disequilibrium (r^2^) of each SNP with the top SNP in the region is indicated by its color. Crossed points represent imputed SNPs, circles represent directly genotyped SNPs.

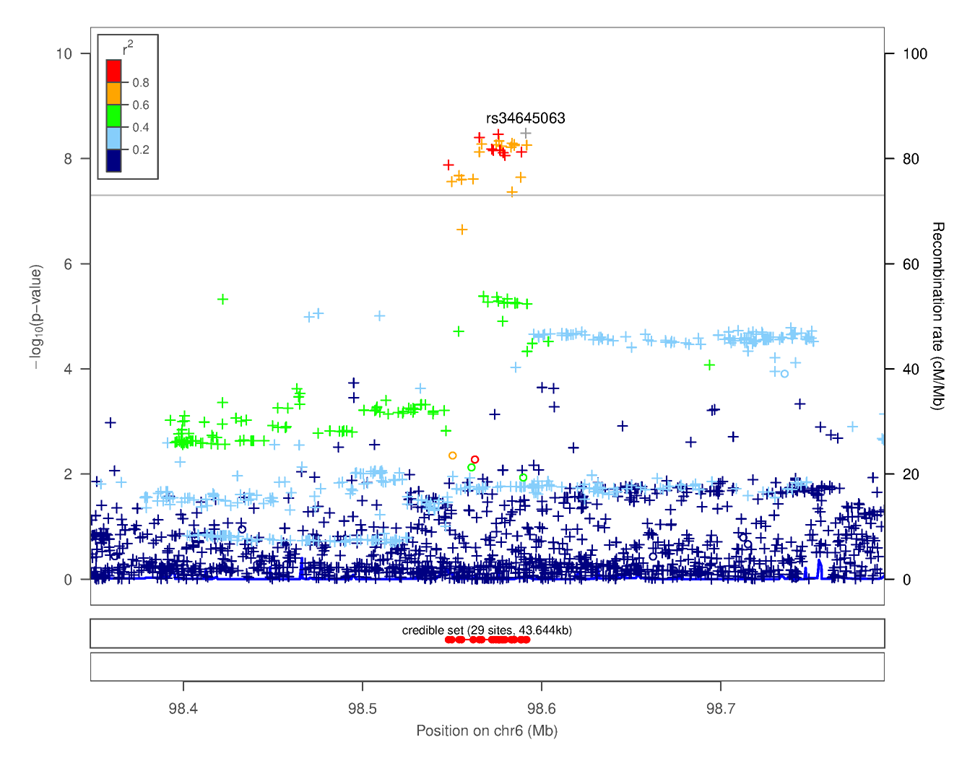

**Supplementary Figure 6.** Locus zoom plot focusing on SNP rs34645063 on chromosome 6. This plot was generated using LocusZoom^7^. The -log_10_(*p-*value) is shown on the left *y*-axis; position in Mb is on the *x*-axis. Recombination rates (expressed in centiMorgans cM per Mb; NCBI Build GRCh37; highlighted in blue) are shown on the right *y*-axis. Pairwise linkage disequilibrium (r^2^) of each SNP with the top SNP in the region is indicated by its color. Crossed points represent imputed SNPs, circles represent directly genotyped SNPs.

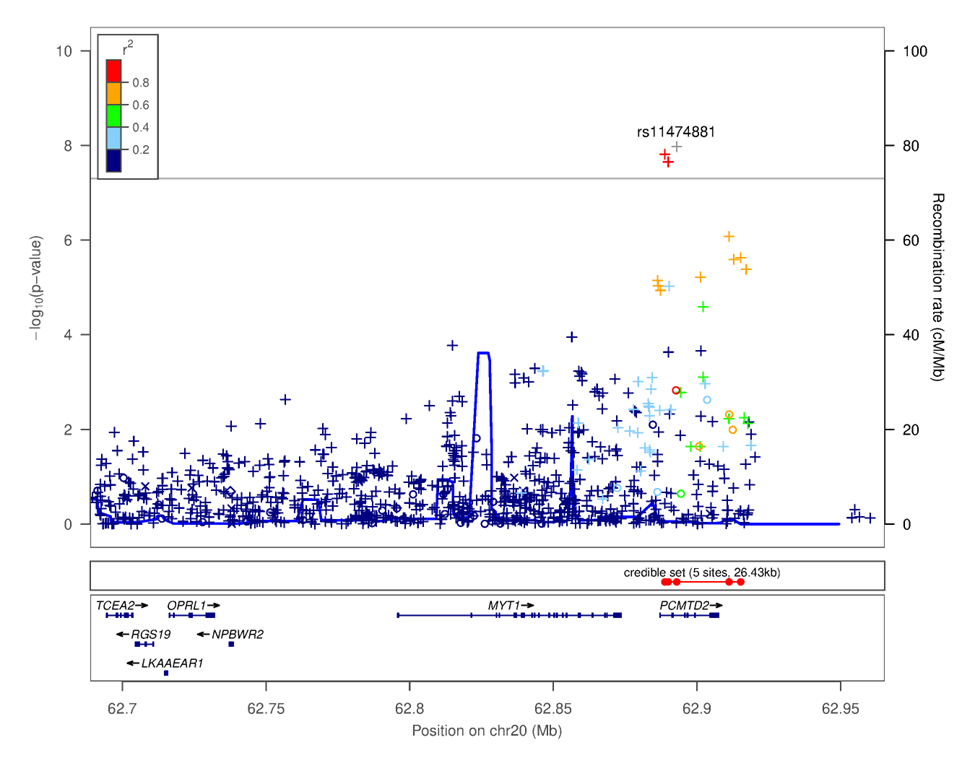

**Supplementary Figure 7.** Locus zoom plot focusing on SNP rs11474881 on chromosome 20. This plot was generated using LocusZoom^7^. The -log_10_(*p-*value) is shown on the left *y*-axis; position in Mb is on the *x*-axis. Recombination rates (expressed in centiMorgans cM per Mb; NCBI Build GRCh37; highlighted in blue) are shown on the right *y*-axis. Pairwise linkage disequilibrium (r^2^) of each SNP with the top SNP in the region is indicated by its color. Crossed points represent imputed SNPs, circles represent directly genotyped SNPs.

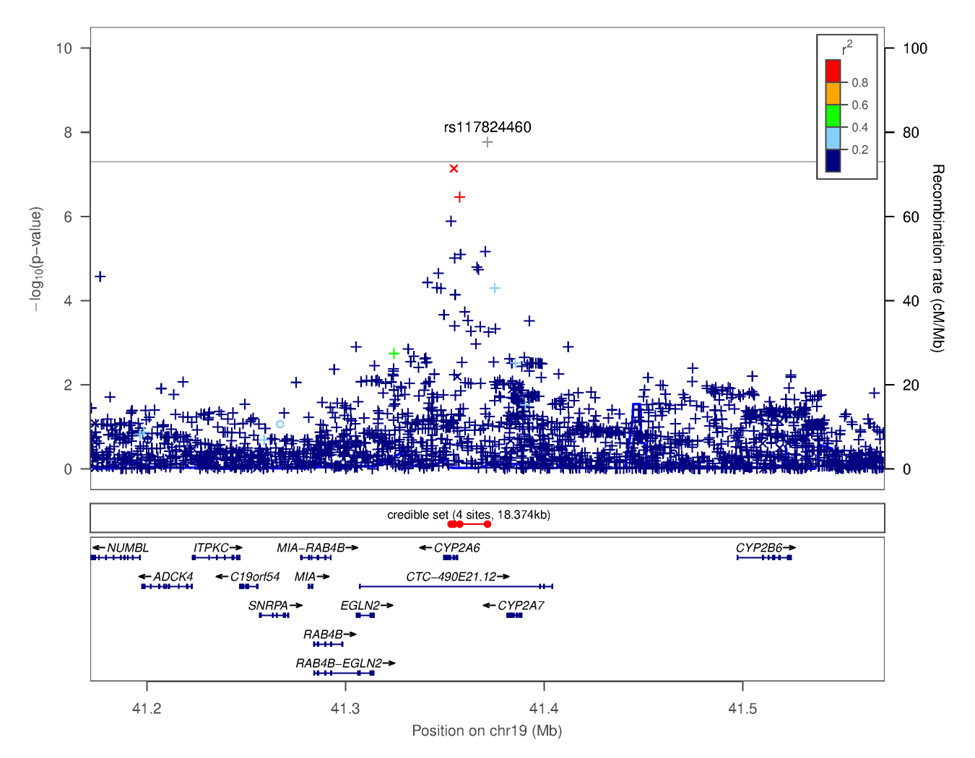

**Supplementary Figure 8.** Locus zoom plot focusing on SNP rs117824460 on chromosome 19. This plot was generated using LocusZoom^7^. The -log_10_(*p-*value) is shown on the left *y*-axis; position in Mb is on the *x*-axis. Recombination rates (expressed in centiMorgans cM per Mb; NCBI Build GRCh37; highlighted in blue) are shown on the right *y*-axis. Pairwise linkage disequilibrium (r^2^) of each SNP with the top SNP in the region is indicated by its color. Crossed points represent imputed SNPs, circles represent directly genotyped SNPs.

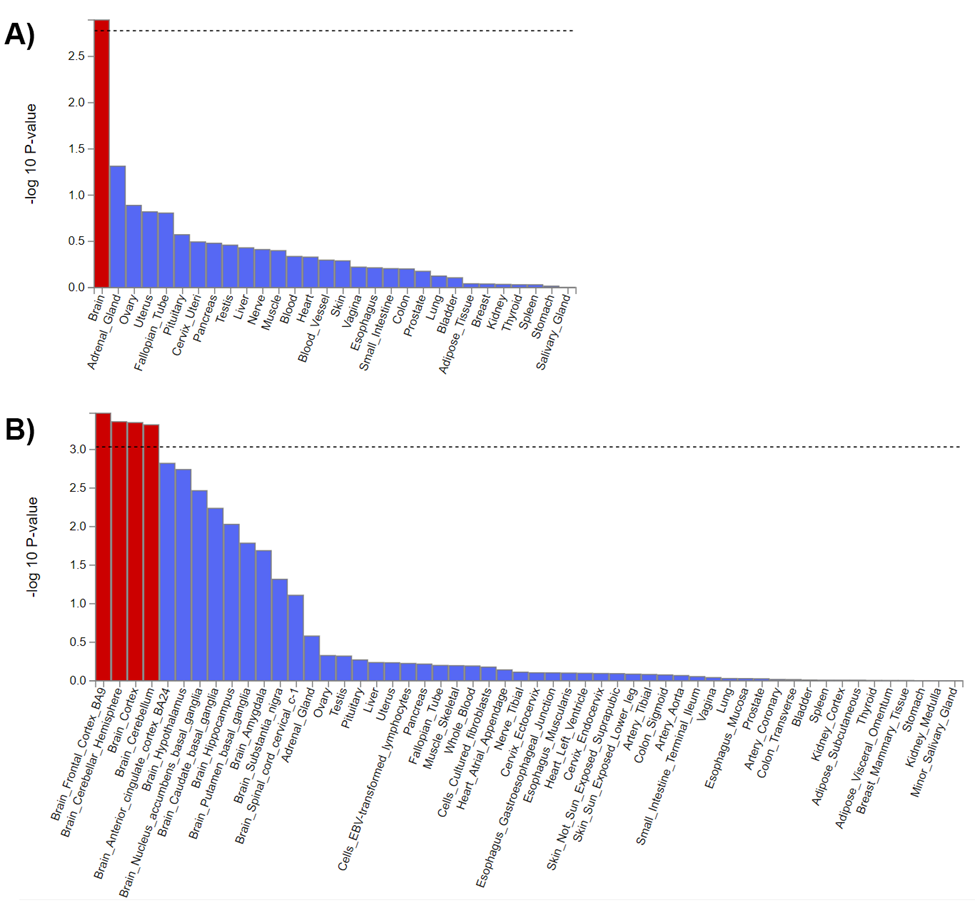

**Supplementary Figure 9.** MAGMA tissue-based enrichment analyses for coffee intake in 23andMe research participants. Analysis conducted across **A)** 30 and **B)** 54 GTEx (v8) tissues with FUMA.

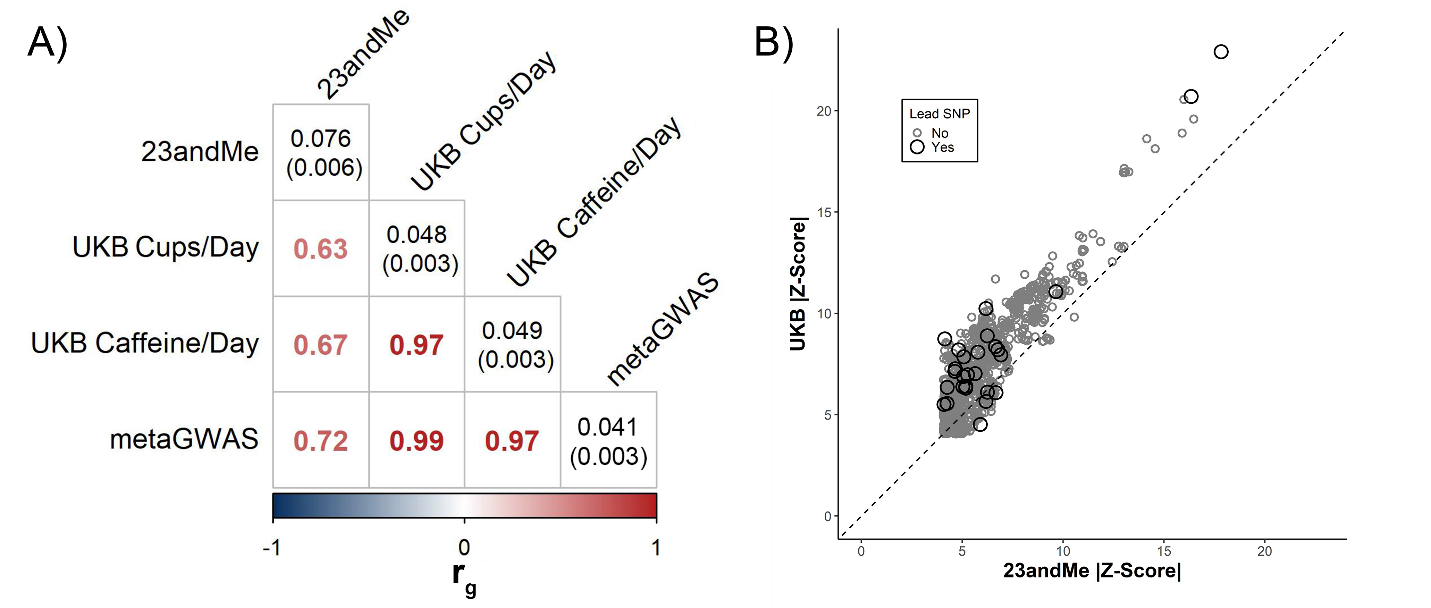

**Supplementary Figure 10.** Meta-analysis of coffee intake summary statistics from the 23andMe (*N=*130,153) and UKB (*N=*334,659) cohorts reveals cohort heterogeneity. **A)** LDSC *r_g_* (± standard error) between summary statistics of coffee intake reveals moderate correlations between 23andMe and UKB data. Heritability estimates (± standard error) presented in the diagonal. metaGWAS refers to the meta-analysis of the 23andMe and UKB coffee cups/day intake summary statistics. UKB Caffeine/Day represents estimates of daily caffeine intake from coffee summary statistics from Said, et al. ^8^. **B)** Comparison of the top SNPs (*p<*5.00E-05) and lead SNPs absolute Z-scores shared between the UKB and 23andMe summary statistics. All top SNPs share the same direction of effect in association with coffee intake.

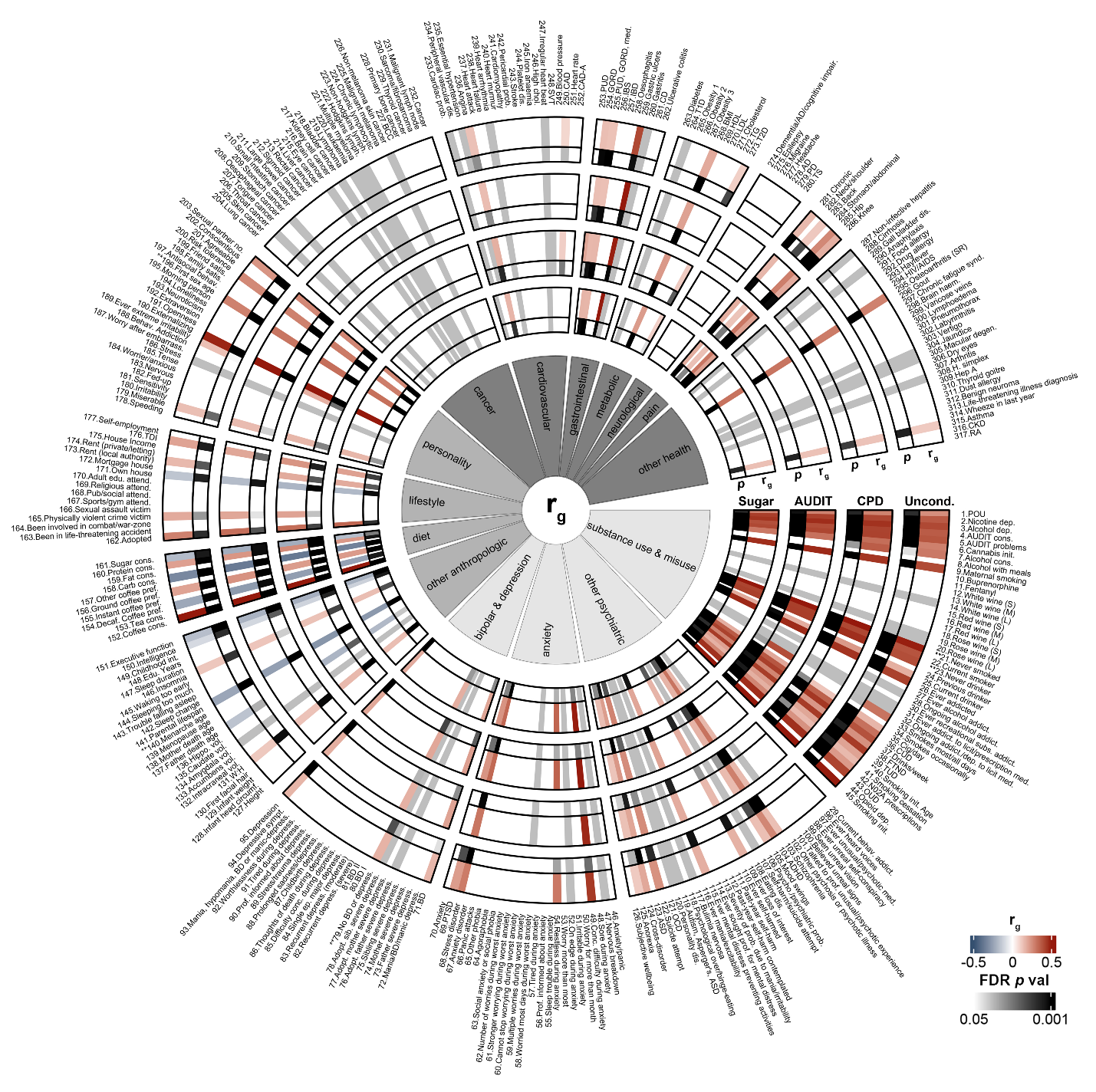

**Supplementary Figure 11.** Genetic correlations with psychiatric (light gray), anthropologic (medium gray), and health (dark gray) 23andMe summary statistics of coffee intake persist following conditional analysis. Figure illustrates unconditioned (lanes 1 and 2), cigarettes per day (CPD; lanes 3 and 4), AUDIT consumption factor (AUDIT; lanes 5 and 6), and dietary sugar intake (Sugar; lanes 7 and 8) genetic correlations across 317 traits. *r_g_* values calculated with LDSC shown in lanes 1, 3, 5, and 7. FDR-corrected *p* values shown in lanes 2, 4, 6, and 8. For a full list of correlations and trait names, see **Supplementary Table 14**. Genetic correlations for traits denoted with * could not be calculated in both cohorts; ** denotes reverse coding. Conditional analysis was conducted using mtCOJO^9^.

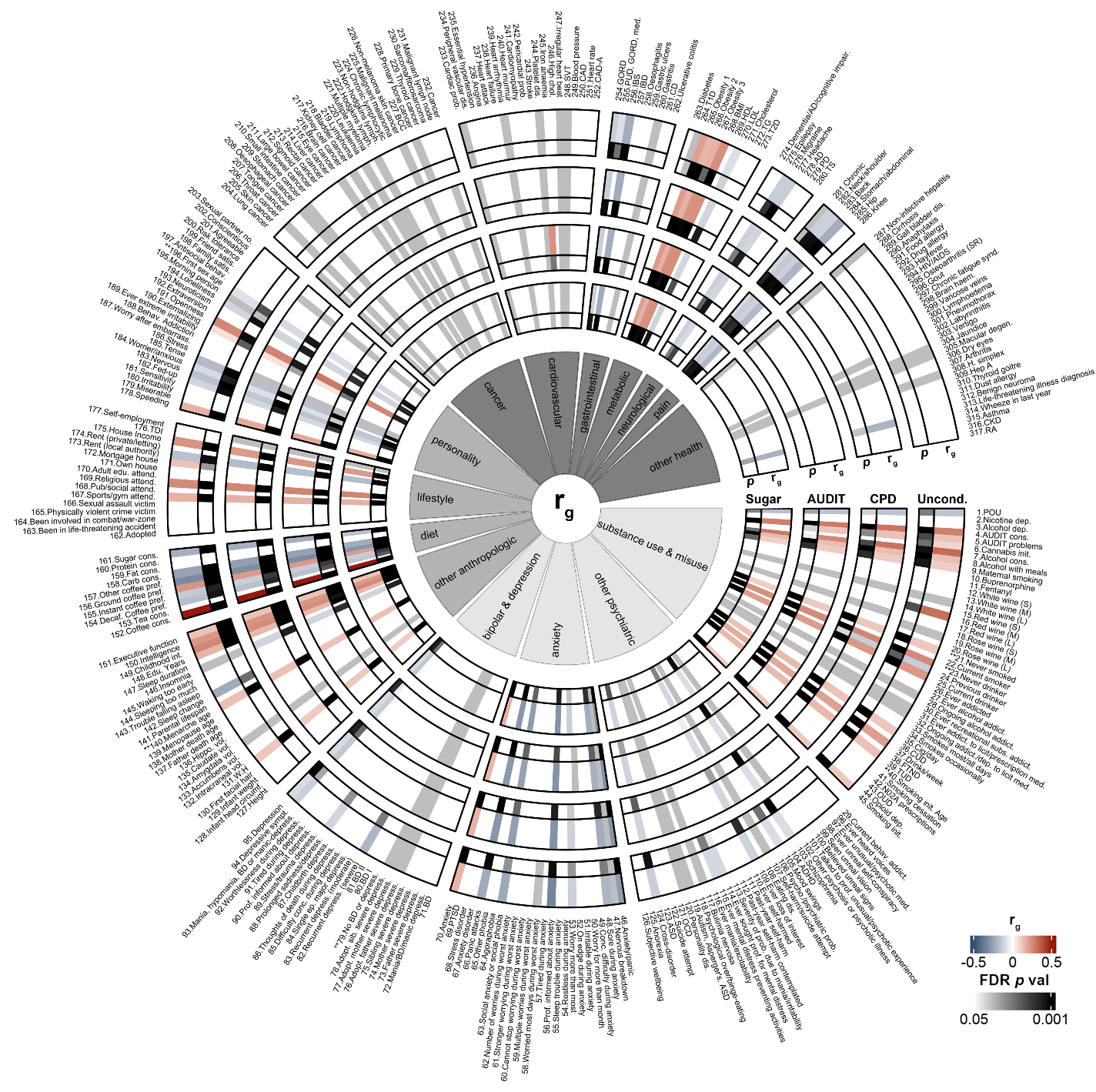

**Supplementary Figure 12.** Genetic correlations with psychiatric (light gray), anthropologic (medium gray), and health (dark gray) UKB summary statistics of coffee intake persist following conditional analysis. Figure illustrates unconditioned (lanes 1 and 2), cigarettes per day (CPD; lanes 3 and 4), AUDIT consumption factor (AUDIT; lanes 5 and 6), and dietary sugar intake (Sugar; lanes 7 and 8) genetic correlations across 317 traits. *r_g_* values calculated with LDSC shown in lanes 1, 3, 5, and 7. FDR-corrected *p* values shown in lanes 2, 4, 6, and 8. For full list of correlations and trait names, see **Supplementary Table 14**. Genetic correlations for traits denoted with * could not be calculated in both cohorts; ** denotes reverse coding. Conditional analysis was conducted using mtCOJO^9^.

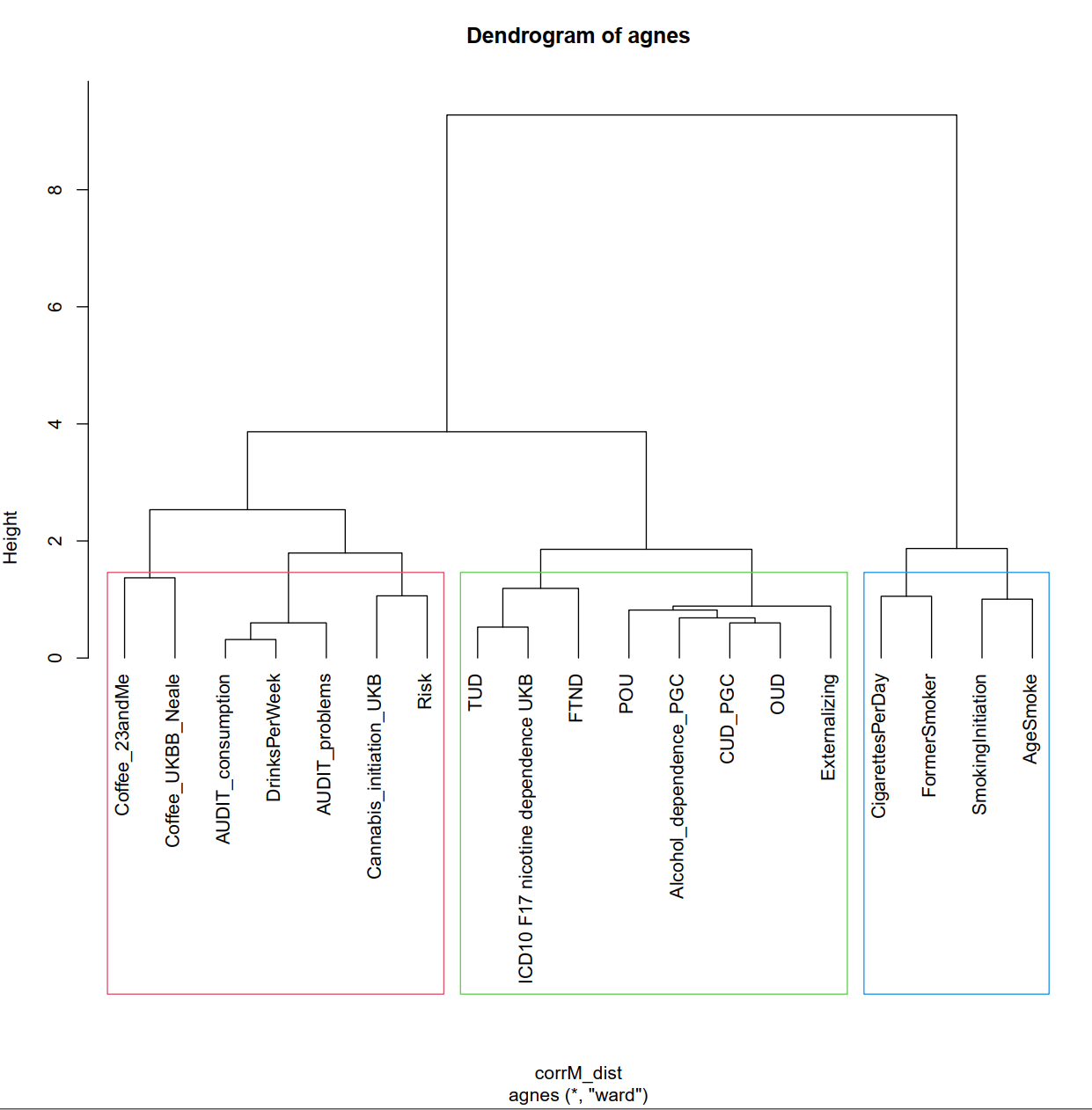

**Supplementary Figure 13.** Cluster analysis reveals that UKB coffee intake is more aligned with summary statistics of substance consumption traits and not substance misuse traits. *Abbreviations.* Coffee_23andMe, Coffee cups/day from 23andMe; Coffee_UKBB_Neale, Coffee cups/day from UKB; TUD, tobacco use disorder; FTND, Fagerström Test for Nicotine Dependence; POU, Problematic Opioid Use; CUD_PGNC, Cannabis Use Disorder; OUD, Opioid Use Disorder (MVP); Risk, General Risk Tolerance; AgeSmoke, Age of Initiation.

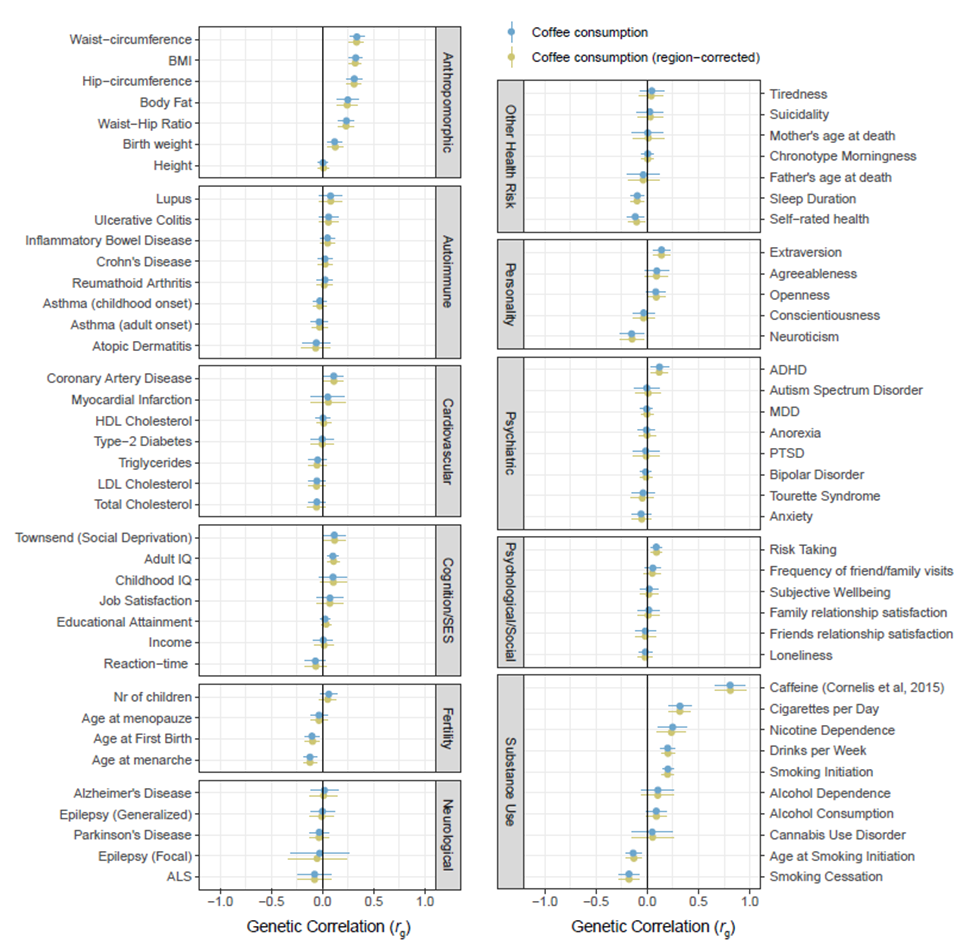

**Supplementary Figure 14.** Controlling for geographic region did not affect genetic correlations with coffee intake in the UKB. For UKB participants for whom coffee intake, birth location, and current location were available (*N=*256,842), coffee intake genetic correlations with health, anthropologic, and psychiatric traits were conditioned on location variables according to Abdellaoui, et al. ^10^.

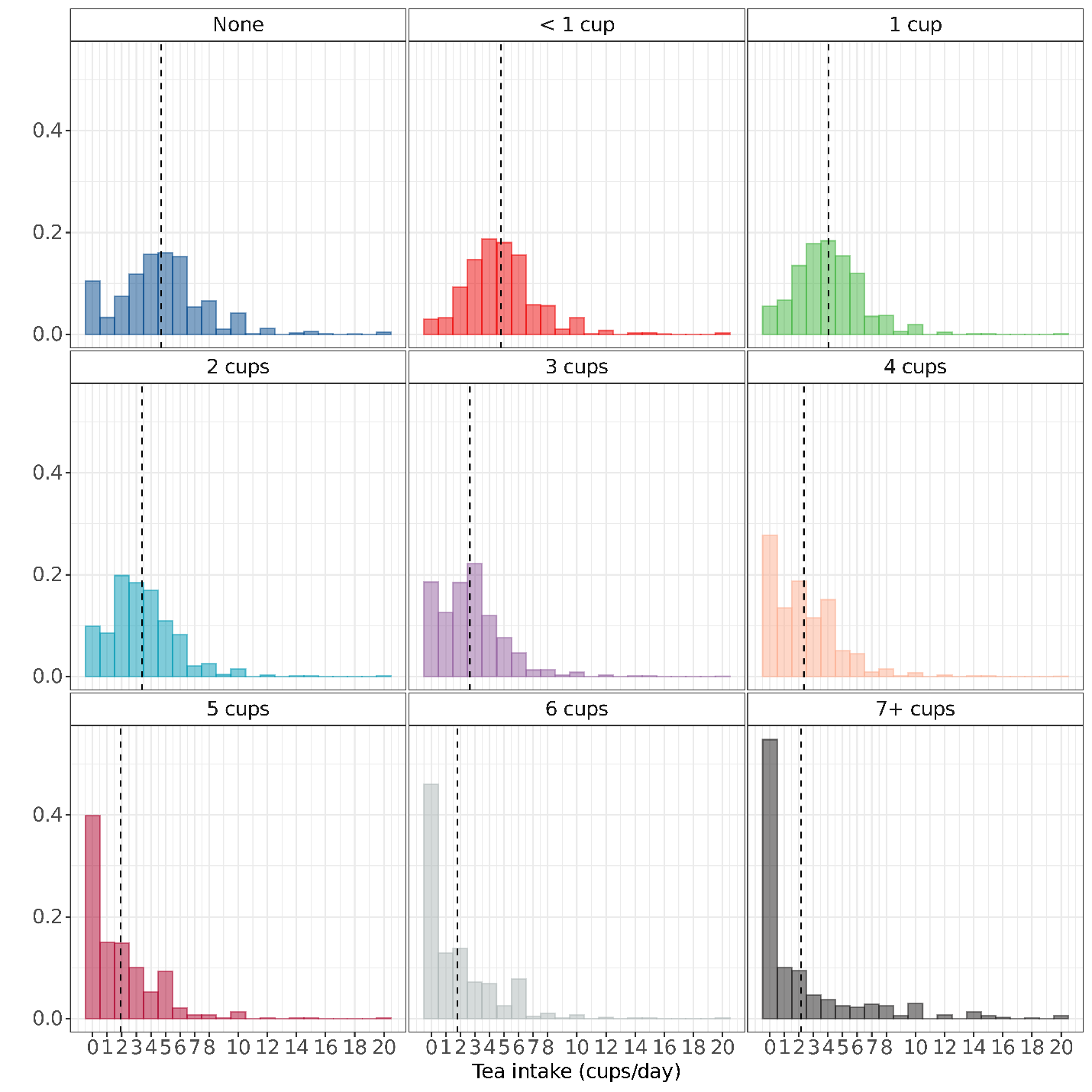

**Supplementary Figure 15.** UKB participants with greater daily coffee intake drink fewer cups of tea and vice versa. Histograms of cups of tea consumed per day categorized by cups of coffee consumed per day in UKB participants (*N=*396,057). Individuals reporting >20 cups of tea per day not shown.

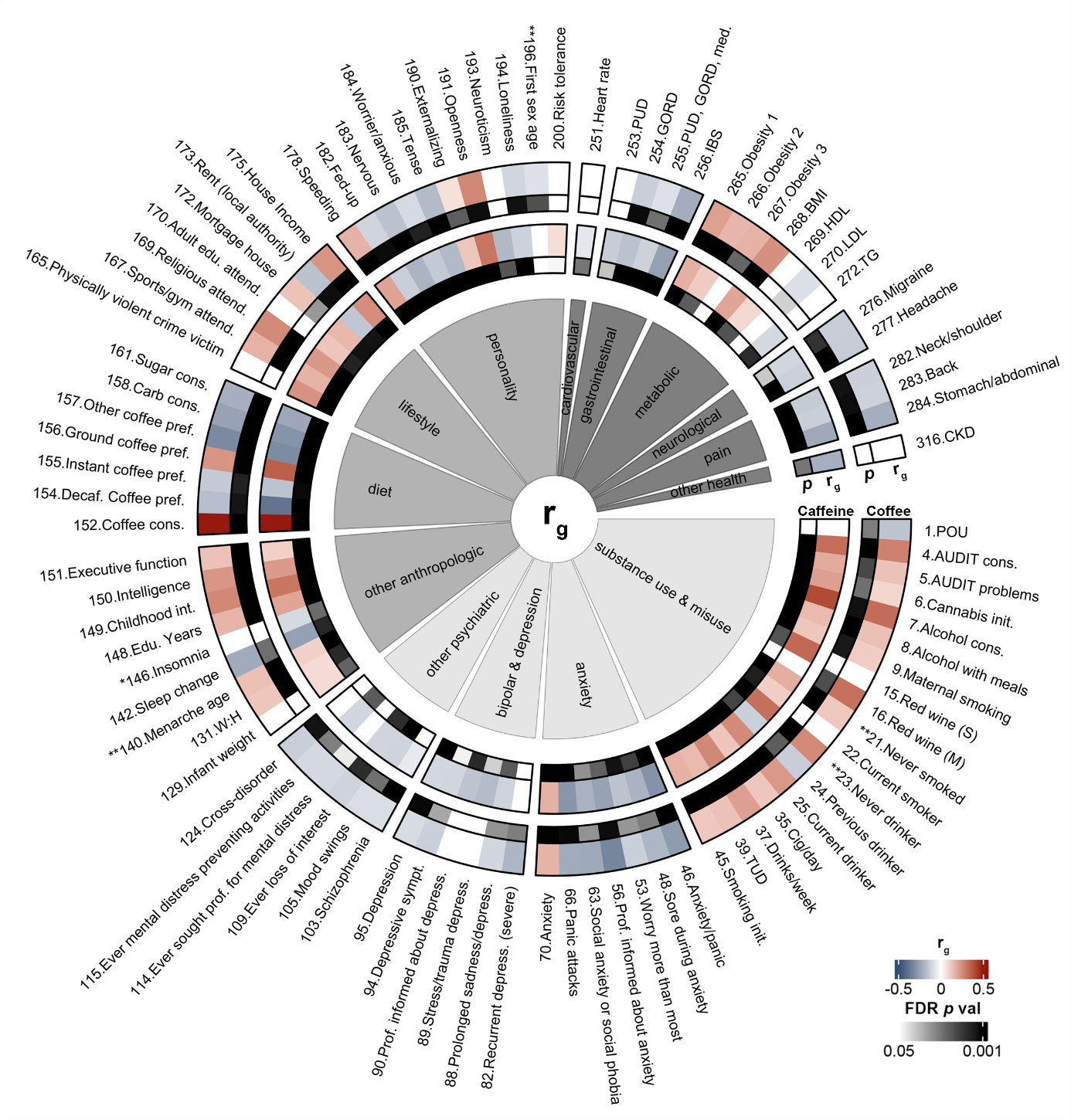

**Supplementary Figure 16.** Comparison of genetic correlations reveals similar results between summary statistics of coffee intake cups per day versus caffeine intake from cups of coffee per day^8^ from the UKB. Figure illustrates coffee intake (Coffee; lanes 1 and 2) and caffeine from coffee intake (Caffeine; lanes 3 and 4) significant genetic correlations. *r_g_* values calculated with LDSC shown in lanes 1 and 3. FDR-corrected p values shown in lanes 2 and 4. 74.16% of traits overlapped in significance and no traits significant in both cohorts showed discordant directions of effect. For full list of correlations and trait names, see **Supplementary Table 14**. Genetic correlations for traits denoted with * could not be calculated in both cohorts; ** denotes reverse coding.

**References**

1	Durand, E. Y., Do, C. B., Mountain, J. L. & Macpherson, J. M. Ancestry Composition: A Novel, Efficient Pipeline for Ancestry Deconvolution. *bioRxiv. [Preprint]*, doi:<https://doi.org/10.1101/010512> (2014).

2	Barkley-Levenson, A. M., Lagarda, F. A. & Palmer, A. A. Glyoxalase 1 (GLO1) Inhibition or Genetic Overexpression Does Not Alter Ethanol's Locomotor Effects: Implications for GLO1 as a Therapeutic Target in Alcohol Use Disorders. *Alcohol Clin Exp Res* **42**, 869-878, doi:10.1111/acer.13623 (2018).

3	Distler, M. G. *et al.* Glyoxalase 1 increases anxiety by reducing GABAA receptor agonist methylglyoxal. *J Clin Invest* **122**, 2306-2315, doi:10.1172/JCI61319 (2012).

4	Dawson, G. R. & Tricklebank, M. D. Use of the elevated plus maze in the search for novel anxiolytic agents. *Trends Pharmacol Sci* **16**, 33-36, doi:10.1016/s0165-6147(00)88973-7 (1995).

5	Henn, B. M. *et al.* Cryptic distant relatives are common in both isolated and cosmopolitan genetic samples. *PLoS One* **7**, e34267, doi:10.1371/journal.pone.0034267 (2012).

6	Fuchsberger, C., Abecasis, G. R. & Hinds, D. A. minimac2: faster genotype imputation. *Bioinformatics* **31**, 782-784, doi:10.1093/bioinformatics/btu704 (2015).

7	Pruim, R. J. *et al.* LocusZoom: regional visualization of genome-wide association scan results. *Bioinformatics* **26**, 2336-2337, doi:10.1093/bioinformatics/btq419 (2010).

8	Said, M. A., van de Vegte, Y. J., Verweij, N. & van der Harst, P. Associations of Observational and Genetically Determined Caffeine Intake With Coronary Artery Disease and Diabetes Mellitus. *J Am Heart Assoc* **9**, e016808, doi:10.1161/JAHA.120.016808 (2020).

9	Zhu, Z. *et al.* Causal associations between risk factors and common diseases inferred from GWAS summary data. *Nat Commun* **9**, 224, doi:10.1038/s41467-017-02317-2 (2018).

10	Abdellaoui, A., Dolan, C. V., Verweij, K. J. H. & Nivard, M. G. Gene-environment correlations across geographic regions affect genome-wide association studies. *Nat Genet* **54**, 1345-1354, doi:10.1038/s41588-022-01158-0 (2022).
